# High variation expected in the pace and burden of SARS-CoV-2 outbreaks across sub-Saharan Africa

**DOI:** 10.1101/2020.07.23.20161208

**Authors:** Benjamin L. Rice, Akshaya Annapragada, Rachel E. Baker, Marjolein Bruijning, Winfred Dotse-Gborgbortsi, Keitly Mensah, Ian F. Miller, Nkengafac Villyen Motaze, Antso Raherinandrasana, Malavika Rajeev, Julio Rakotonirina, Tanjona Ramiadantsoa, Fidisoa Rasambainarivo, Weiyu Yu, Bryan T. Grenfell, Andrew J. Tatem, C. Jessica E. Metcalf

## Abstract

A surprising feature of the SARS-CoV-2 pandemic to date is the low burdens reported in sub-Saharan Africa (SSA) countries relative to other global regions. Potential explanations (e.g., warmer environments^1^, younger populations^2–4^) have yet to be framed within a comprehensive analysis accounting for factors that may offset the effects of climate and demography. Here, we synthesize factors hypothesized to shape the pace of this pandemic and its burden as it moves across SSA, encompassing demographic, comorbidity, climatic, healthcare and intervention capacity, and human mobility dimensions of risk. We find large scale diversity in probable drivers, such that outcomes are likely to be highly variable among SSA countries. While simulation shows that extensive climatic variation among SSA population centers has little effect on early outbreak trajectories, heterogeneity in connectivity is likely to play a large role in shaping the pace of viral spread. The prolonged, asynchronous outbreaks expected in weakly connected settings may result in extended stress to health systems. In addition, the observed variability in comorbidities and access to care will likely modulate the severity of infection: We show that even small shifts in the infection fatality ratio towards younger ages, which are likely in high risk settings, can eliminate the protective effect of younger populations. We highlight countries with elevated risk of ‘slow pace’, high burden outbreaks. Empirical data on the spatial extent of outbreaks within SSA countries, their patterns in severity over age, and the relationship between epidemic pace and health system disruptions are urgently needed to guide efforts to mitigate the high burden scenarios explored here.

---

The trajectory of the SARS-CoV-2 pandemic in lower latitude, lower income countries including in Sub-Saharan Africa (SSA) remains uncertain. To date, reported case counts and mortality in SSA have lagged behind other geographic regions: all SSA countries, with the exception of South Africa, reported less than 27,000 total cases as of June 2020 ^5^ (**Table S1**) - totals far less than observed in Asia, Europe, and the Americas ^5,6^. However, recent increases in reported cases in many SSA countries make it unclear whether the relatively few reported cases to date indicate a reduced epidemic potential or rather an initial delay relative to other regions.

Correlation between surveillance capacity and case counts ^7^ obscure early trends in SSA (**Figure S1**). Experience from locations in which the pandemic has progressed more rapidly provides a basis of knowledge to assess the relative risk of populations in SSA and identify those at greatest risk. For example, individuals in lower socio-economic settings have been disproportionately affected in high latitude countries,^8,9^ indicating poverty as an important determinant of risk. Widespread disruptions to routine health services have been reported ^10–12^ and are likely to be an important contributor to the burden of the pandemic in SSA ^13^. The role of other factors from demography ^2–4^ to health system context ^14^ and intervention timing ^15,16^ is also increasingly well-characterized.

## Factors expected to increase and decrease SARS-CoV-2 risk in SSA

Anticipating the trajectory of ongoing outbreaks in SSA requires considering variability in known drivers, and how they may interact to increase or decrease risk across populations in SSA and relative to non-SSA settings (**Figure 1**). For example, while most countries in SSA have ‘young’ populations, suggesting a decreased burden (since SARS-CoV-2 morbidity and mortality increase with age ^2–4^), prevalent infectious and non-communicable comorbidities may counterbalance this demographic ‘advantage’ ^14,17–19^. Similarly, SSA countries have health systems that vary greatly in their infrastructure, and dense, resource-limited urban populations may have fewer options for social distancing ^20^. Yet, decentralized, community-based health systems that benefit from recent experience with epidemic response (e.g., to Ebola ^21,22^) can be mobilized. Climate is frequently invoked as a potential mitigating factor for warmer and wetter settings ^1^, including SSA, but climate varies greatly between population centers in SSA and large susceptible populations may counteract any climate forcing during initial phases of the epidemic ^23^. Connectivity, at international and subnational scales, also varies greatly ^24,25^ and the time interval between viral introductions and the onset of interventions such as lockdowns will modulate the trajectory ^7^. Finally, burdens of malnutrition, infectious diseases, and many other underlying health conditions are higher in SSA (**Table S2**), and their interactions with SARS-CoV-2 are, as of yet, poorly understood.

**Figure 1.**
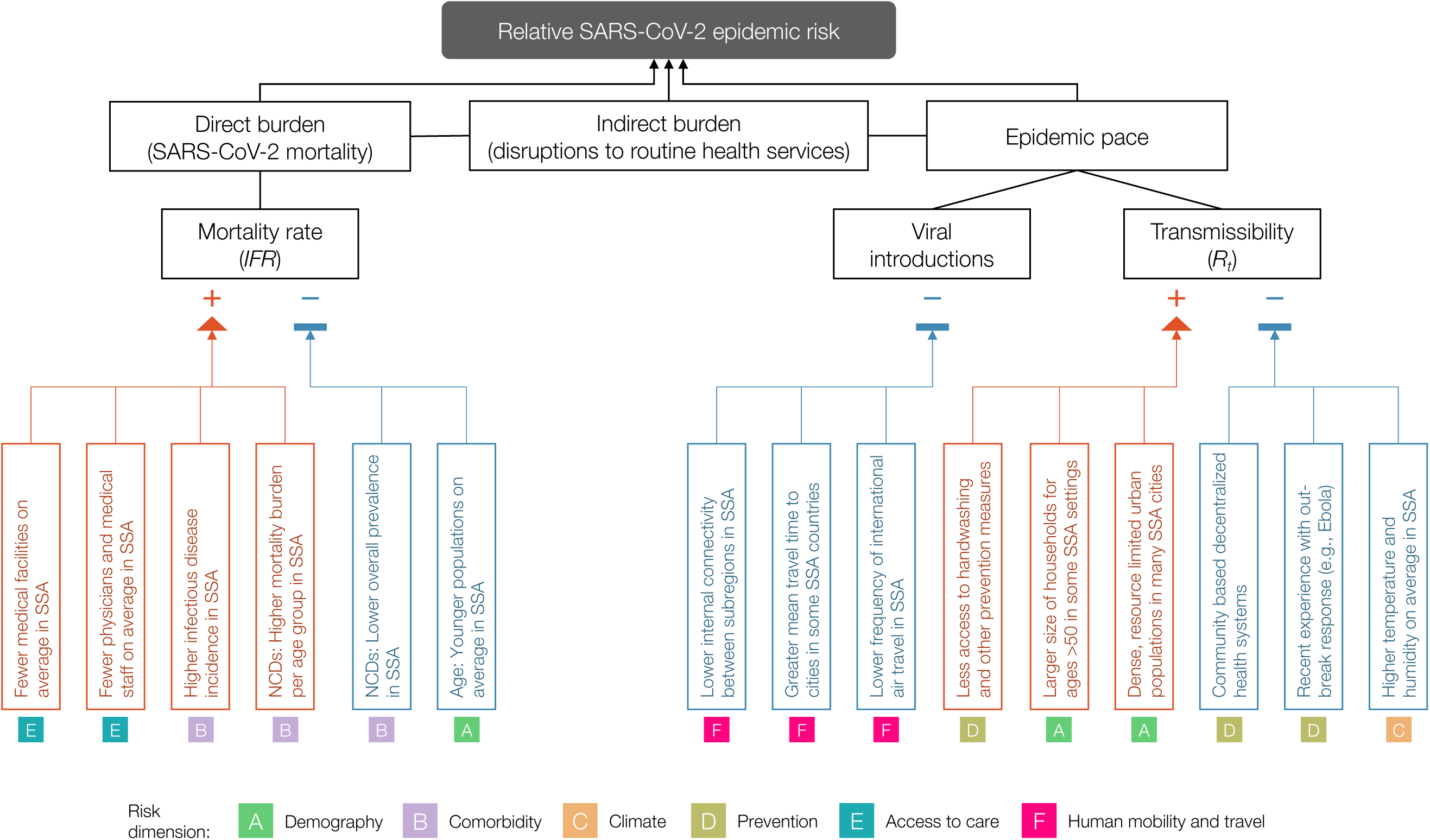
Hypothesized modulators of relative SARS-CoV-2 epidemic risk in sub-Saharan Africa. Factors hypothesized to increase (red) or decrease (blue) mortality burden or epidemic pace within sub-Saharan Africa, relative to global averages, are grouped in six categories or dimensions of risk (A-F). In this framework, epidemic pace is determined by person to person transmissibility (which can be defined as the time-varying effective reproductive number, *R*_*t*_) and introduction and geographic spread of the virus via human mobility. SARS-CoV-2 mortality (determined by the infection fatality ratio, *IFR*) is modulated by demography, comorbidities (e.g., non-communicable diseases (NCDs)), and access to care. Overall burden is a function of direct burden and indirect effects due to, for example, disruptions in health services such as vaccination and infectious disease control. **Table S2** contains details and the references used as a basis to draw the hypothesized modulating pathways.

The highly variable social and health contexts of countries in SSA will drive location-specific variation in the magnitude of the burden, the time-course of the outbreak, and options for mitigation. Here, we synthesize the range of factors hypothesized to modulate the potential outcomes of SARS-CoV-2 outbreaks in SSA settings by leveraging existing data sources and integrating novel SARS-CoV-2 relevant mobility and climate-transmission models. Data on direct measures and indirect indicators of risk factors were sourced from publicly available databases including from the WHO, World Bank, UNPOP, DHS, GBD, and WorldPop, and newly generated data sets (see **Table S3** for details). We organize our assessment around two aspects that will shape national outcomes and response priorities in the event of widespread outbreaks: i) the burden, or expected severity of the outcome of an infection, which emerges from age, comorbidities, and health systems functioning, and ii) the rate of spread within a geographic area, or pace of the pandemic.

We group factors that may drive the relative rates of these two features (mortality burden and pace of the outbreak) along six dimensions of risk: (A) Demographic and socio-economic parameters related to transmission and burden, (B) Comorbidities relevant to burden, (C) Climatic variables that may impact the magnitude and seasonality of transmission, (D) Capacity to deploy prevention measures to reduce transmission, (E) Accessibility and coverage of existing healthcare systems to reduce burden, and (F) Patterns of human mobility relevant to transmission (**Table S2**).

## National and subnational variability in SSA

National scale variability in SSA among these dimensions of risk often exceeds ranges observed across the globe (**Figure 2A-D**). For example, estimates of access to basic handwashing (i.e., clean water and soap ^26^) among urban households in Mali, Madagascar, Tanzania, and Namibia (62-70%) exceed the global average (58%), but fall to less than 10% for Liberia, Lesotho, Congo DRC, and Guinea-Bissau (**Figure 2D**). Conversely, the range in the number of physicians is low in SSA, with all countries other than Mauritius below the global average (168.78 per 100,000 population) (**Figure 2A**). Yet, estimates are still heterogeneous within SSA, with, for example, Gabon estimated to have more than 4 times the physicians of neighboring Cameroon (36.11 and 8.98 per 100,000 population, respectively). This disparity is likely to interact with social contact rates among the elderly in determining exposure and clinical outcomes (e.g., for variation in household size see **Figure 2E-F**). Relative ranking across variables is also uneven among countries with the result that this diversity cannot be easily reduced (e.g., the first two principal components explain only 32.6%, and 13.1% of the total variance as shown in **Figure S5**), motivating a more holistic approach to projecting burden.

**Figure 2.**
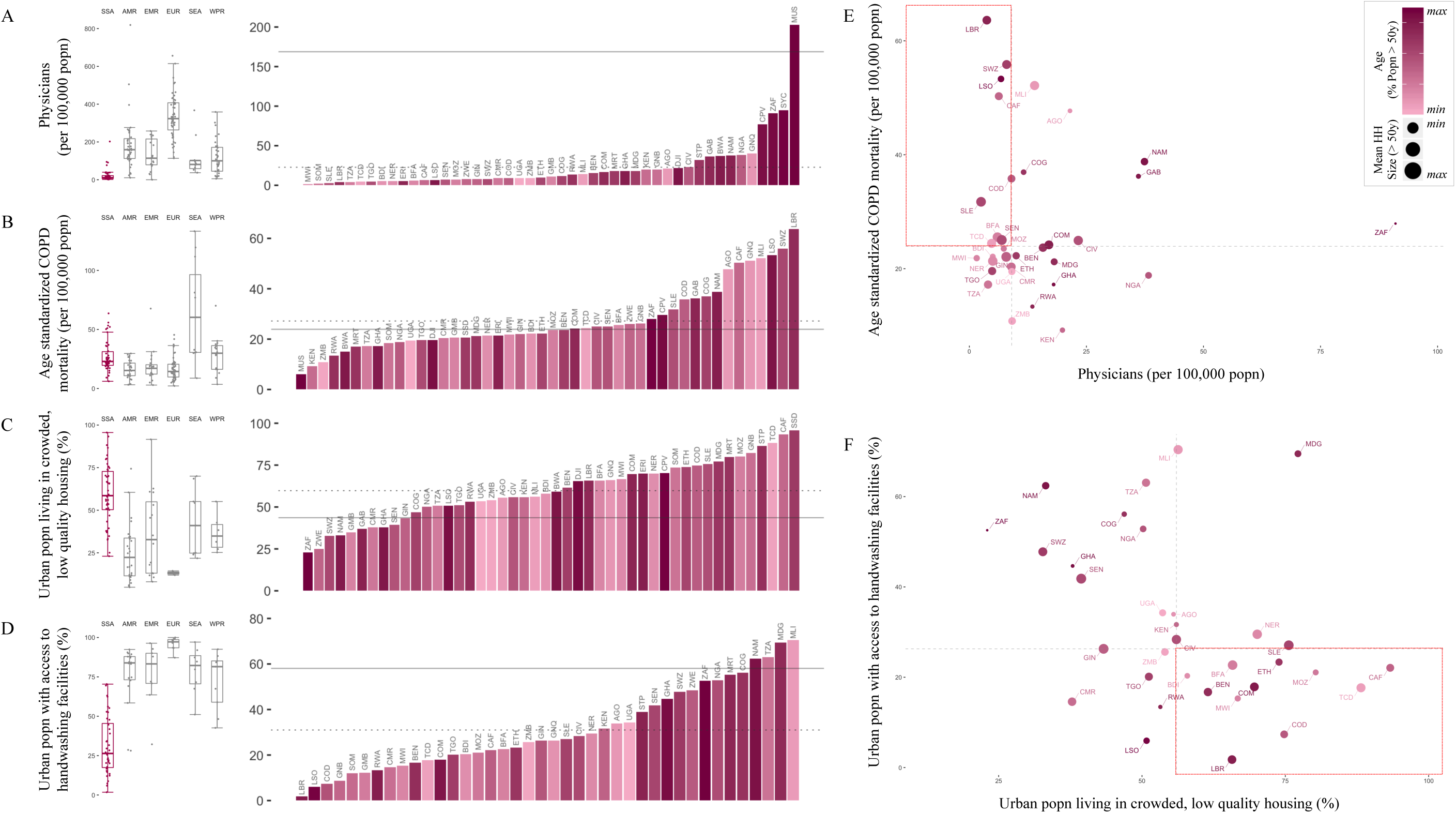
Variation among sub-Saharan African countries in select determinants of SARS-CoV-2 risk. **A-D:** At right, SSA countries are ranked from least to greatest for each indicator; bar color shows population age structure (% of the population above age 50). Solid horizontal lines show the global mean value; dotted lines show the mean among SSA countries. At left, boxplots show median and interquartile range, grouped by geographic region, per WHO: sub-Saharan Africa (SSA); Americas Region (AMR); Eastern Mediterranean Region (EMR); Europe Region (EUR); Southeast Asia Region (SEA); Western Pacific Region (WPR). **E-F:** Dot size shows mean household (HH) size for HHs with individuals over age 50; dashed lines show median value among SSA countries; quadrants of greatest risk are outlined in red (e.g., fewer physicians and greater age standardized Chronic Obstructive Pulmonary Disease (COPD) mortality). See Table S3, Figure S3, and the [SSA-SARS-CoV-2-tool] for full description and visualization of all variables.

## Severity of infection outcome

To first evaluate variation in the burden emerging from the severity of infection outcome, we consider how demography, comorbidity, and access to care might modulate the age profile of SARS-CoV-2 morbidity and mortality ^2–4^. Subnational variation in the distribution of high risk age groups indicates considerable variability, with higher burden expected in urban settings in SSA (**Figure 3A**), where density and thus transmission are likely higher ^27^.

**Figure 3.**
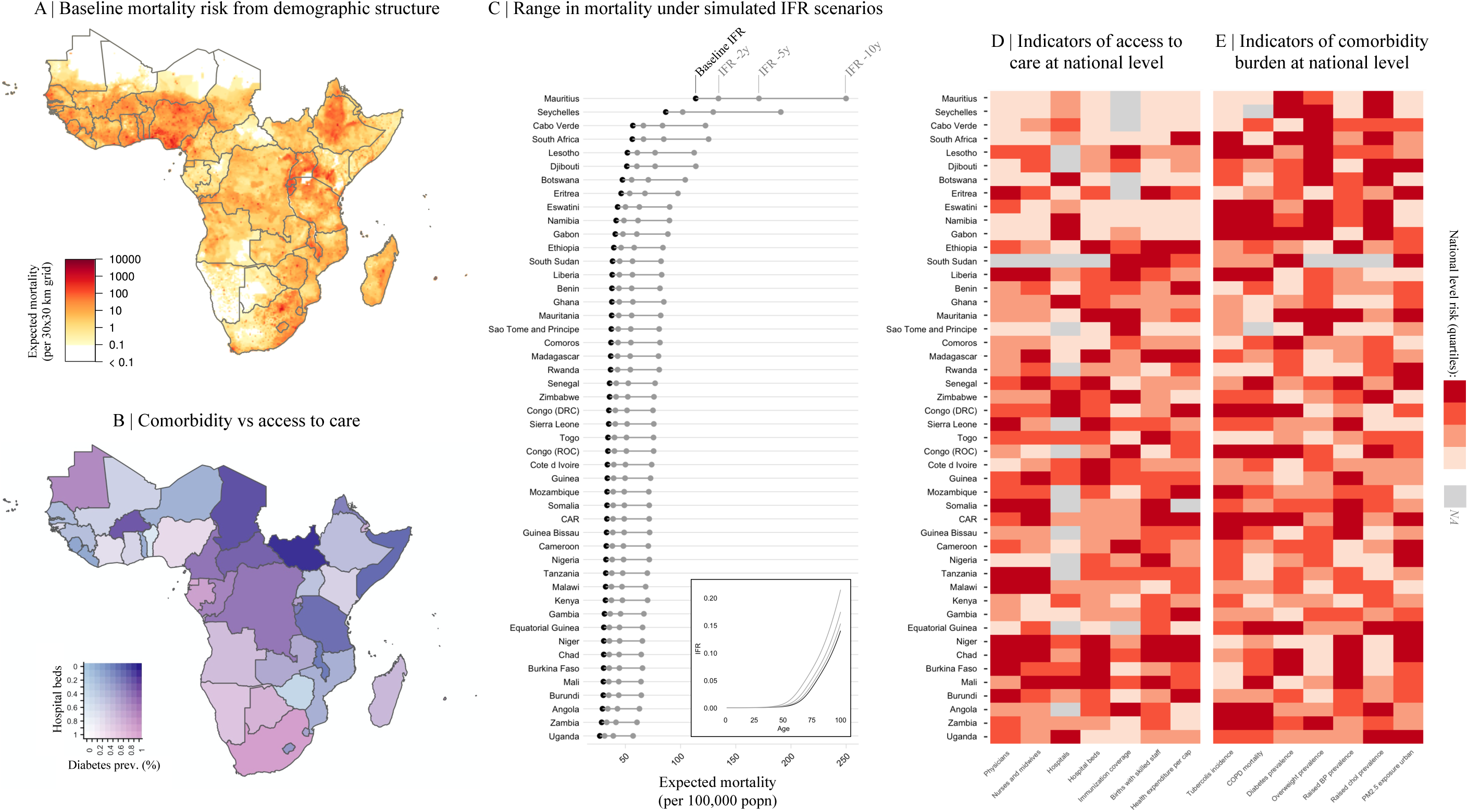
Variation in expected burden for SARS-CoV-2 outbreaks in sub-Saharan Africa. **A:** Expected mortality in a scenario where cumulative infection reaches 20% across age groups and the infection fatality ratio (*IFR*) curve is fit to existing age-stratified *IFR* estimates (see methods, **Table S4**). **B**: National level variation in comorbidity and access to care variables, for e.g., diabetes prevalence among adults and the number of hospital beds per 100,000 population for sub-Saharan African countries. **C**: The range in mortality per 100,000 population expected in scenarios where cumulative infection rate is 20% and *IFR* per age is the baseline (black) or shifted 2, 5, or 10 years younger (gray). Inset, the *IFR* by age curves for each scenario. **D-E**: Select national level indicators; estimates of reduced access to care (e.g., fewer hospitals) or increased comorbidity burden (e.g., higher prevalence of raised blood pressure) shown with darker red for higher risk quartiles (see **Figure S4** for all indicators). Countries missing data for an indicator (NA) are shown in gray. For comparison between countries, estimates are age-standardized where applicable (see **Table S3** for details). See the [SSA-SARS-CoV-2-tool] for high resolution maps for each variable and scenario.

Comorbidities and access to clinical care also vary across SSA (e.g., for diabetes prevalence and hospital bed capacity see **Figure 3B**). In comparison to settings where previous SARS-CoV-2 infection fatality ratio (*IFR*) estimates have been reported, mortality due to noncommunicable diseases in SSA increases more rapidly with age (**Figure S6**). Consequently, we explore scenarios where the SARS-CoV-2 *IFR* increases more rapidly with age than the baseline expected from other settings. Small shifts (e.g., of 2-10 years) in the *IFR* profile result in large effects on expected mortality for a given level of infection. For example, Chad, Burkina Faso, and the Central African Republic, while among the youngest SSA countries, have a relatively high prevalence of diabetes and relatively low density of hospital beds. A five year shift younger in the *IFR* by age profile of SARS-CoV-2 in these settings would result in nearly a doubling of mortality, to a rate that would exceed the majority of other, ‘older’ SSA countries at the unshifted baseline (**Figure 3C**, see supplement for details of methods). Although there is greater access to care in older populations by some metrics (**Figure 2A**, correlation between age and the number of physicians per capita, *r* = 0.896, *p* < 0.001), access to clinical care is highly variable overall (**Figure 3D**) and maps poorly to indicators of comorbidity (**Figure 3E**). Empirical data are urgently needed to assess the extent to which the *IFR*-age-comorbidity associations observed elsewhere are applicable to SSA settings with reduced access to advanced care. Yet both surveillance and mortality registration ^28^ are frequently under-resourced in SSA, complicating both evaluating and anticipating the burden of the pandemic, and underscoring the urgency of strengthening existing systems ^22^.

## Pandemic pace

Next, we turn to the pace of the pandemic within each country. The frequency of viral introduction to each country, likely governed by international air travel in SSA ^29^, determines both the timing of the first infections and the number of initial infection clusters that can seed subsequent outbreaks. The relative importation risk among SSA cities and countries was assessed by compiling data from 108,894 flights arriving at 113 international airports in SSA from January to April 2020 (**Figure 4A**), stratified by the SARS-CoV-2 status at the departure location on the day of travel (**Figure 4B**). A small subset of SSA countries received a disproportionately large percentage (e.g., South Africa, Ethiopia, Kenya, Nigeria together contribute 47.9%) of the total travel from countries with confirmed SARS-CoV-2 infections, likely contributing to variation in the pace of the pandemic across settings ^29,30^.

**Figure 4.**
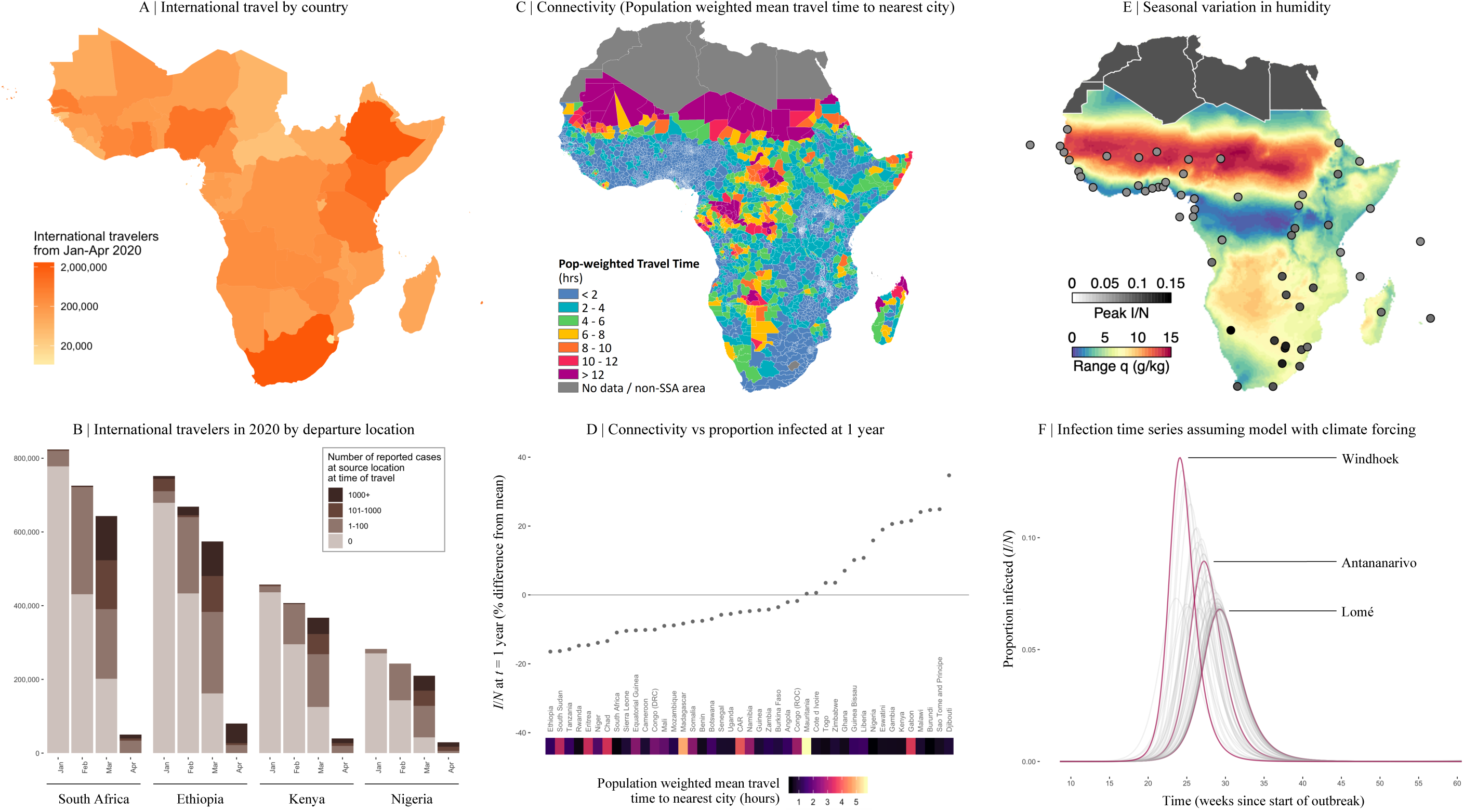
Variation in connectivity and climate in sub-Saharan Africa and expected effects on SARS-CoV-2. **A:** International travellers to sub-Saharan Africa (SSA) from January to April 2020, as inferred from the number of passenger seats on arriving aircraft. **B**: For the four countries with the most arrivals, the proportion of arrivals by month coming from countries with 0, 1-100, 101-1000, and 1000+ reported SARS-CoV-2 infections at the time of travel (see **Table S5** for all others). **C**: Connectivity within SSA countries as inferred from average population weighted mean travel time to the nearest urban area greater than 50,000 population. **D**: Mean travel time at the national level and variation in the fraction of the population expected to be infected (*I/N*) in the first year from stochastic simulations (see methods). **E**: Climate variation across SSA as shown by seasonal range in specific humidity, *q* (g/kg) (max average *q* - min average *q*). Circles show peak proportion infected. **F:** The effect of local seasonality in SSA cities on outbreaks (*I/N* over time) in susceptible populations beginning in March 2020 (see methods).

Once local chains of infection are established, the rate of spread within countries will be shaped by efforts to reduce spread, such as handwashing (**Figure 2D**), population contact patterns including mobility and urban crowding ^27^ (e.g., **Figure 2C**), and potentially the effect of climatic variation ^1^. Where countries fall across this spectrum of pace will shape interactions with lockdowns and determine the length and severity of disruptions to routine health system functioning.

Subnational connectivity varies greatly across SSA, both between subregions of a country and between cities and their rural periphery (e.g., as indicated by travel time to the nearest city over 50,000 population, **Figure 4C**). As expected, in stochastic simulations using estimates of viral transmission parameters and mobility (assuming no variation in control efforts, see methods), a smaller cumulative proportion of the population is infected at a given time in countries with larger populations in less connected subregions (**Figure 4D**). At the national level, susceptibility declines more slowly and more unevenly in such settings (e.g., Ethiopia, South Sudan, Tanzania) due to a lower probability of introductions and re-introductions of the virus locally; an effect amplified by lockdowns. It remains unclear whether the more prolonged, asynchronous epidemics expected in these countries or the overlapping, concurrent epidemics expected in countries with higher connectivity (e.g. Malawi, Kenya, Burundi) will be a greater stress to health systems. Outbreak control efforts are likely to be further complicated during prolonged epidemics if they intersect with seasonal events such as temporal patterns in human mobility ^31^ or other infections (e.g., malaria).

Turning to climate, despite extreme variation among cities in SSA (**Figure 4E**), large epidemic peaks are expected in all cities (**Figure 4F**), even from models where transmission rate significantly declines in warmer, more humid settings. In the absence of interventions, with transmission rate modified by climate only, peak timing varies only by 4-6 weeks with peaks generally expected earlier in more southerly, colder, drier, cities (e.g., Windhoek and Maseru) and later in more humid, coastal cities (e.g., Bissau, Lomé, and Lagos). Apart from these slight shifts in timing, large susceptible populations overwhelm the effects of climate ^23^, and earlier suggestions that Africa’s generally more tropical environment may provide a protective effect^1^ are not supported by evidence.

## Context-specific preparedness in SSA

Our synthesis emphasizes striking country to country variation in drivers of the pandemic in SSA (**Figure 2**), indicating variation in the burden (**Figure 3**) and pace (**Figure 4**) is to be expected even across low income settings. As small perturbations in the age profile of mortality could drastically change the national level burden in SSA (**Figure 3**), building expectations for the risk for each country requires monitoring for deviations in the pattern of morbidity and mortality over age. Transparent and timely communication of these context-specific risk patterns could help motivate population behavioral changes and guide existing networks of community case management.

Because the largest impacts of SARS-CoV-2 outbreaks may be through indirect effects on routine health provisioning, understanding how existing programs may be disrupted differently by acute versus longer outbreaks is crucial to planning resource allocation. For example, population immunity will decline proportionally with the length of disruptions to routine vaccination programs ^31^, resulting in more severe consequences in areas with prolonged epidemic time courses.

Others have suggested that this crisis presents an opportunity to unify and mobilize across existing health programs (e.g., for HIV, TB, Malaria, and other NCDs) ^22^. While this may be a powerful strategy in the context of acute, temporally confined crises, long term distraction and diversion of resources ^32^ may be harmful in settings with extended, asynchronous epidemics. A higher risk of infection among healthcare workers during epidemics ^33,34^ may amplify this risk. Due to the lag relative to other geographic regions, many SSA settings retain the opportunity to prepare for and intervene in the earlier epidemic phases via context-specific deployment of both routine and pandemic related interventions. As evidenced by failures in locations where the epidemic progressed rapidly (e.g., USA), effective governance and management prior to reaching large case counts is likely to yield the largest rewards. Mauritius ^35^ and Rwanda ^36^, for example, have reported extremely low incidence thanks in part to a well-managed early response.

## Conclusions

The burden and time-course of SARS-CoV-2 is expected to be highly variable across sub-Saharan Africa. As the outbreak continues to unfold, critically evaluating this mapping to better understand where countries lie in terms of their relative risk (e.g., see **Figure 5**) will require increased surveillance, and timely documentation of morbidity and mortality over age. Case counts are rising across SSA, but variability in testing regimes makes it difficult to compare observations to date with expectations in terms of pace (**Figure S7**). The potential to miss large clusters of cases (in contexts with weaker surveillance), combined with the potential that large areas remain unreached by the pandemic for longer (as a result of slower ‘pace’), indicate that immunological surveys are likely a powerful lens for understanding the landscape of population risk ^37^. When considering hopeful futures with the possibility of a SARS-CoV-2 vaccine, it is imperative that vaccine distribution be equitable, and in proportion with need. Understanding factors that both drive spatial variation in vulnerable populations and temporal variation in pandemic progression could help approach these goals in SSA.

**Figure 5.**
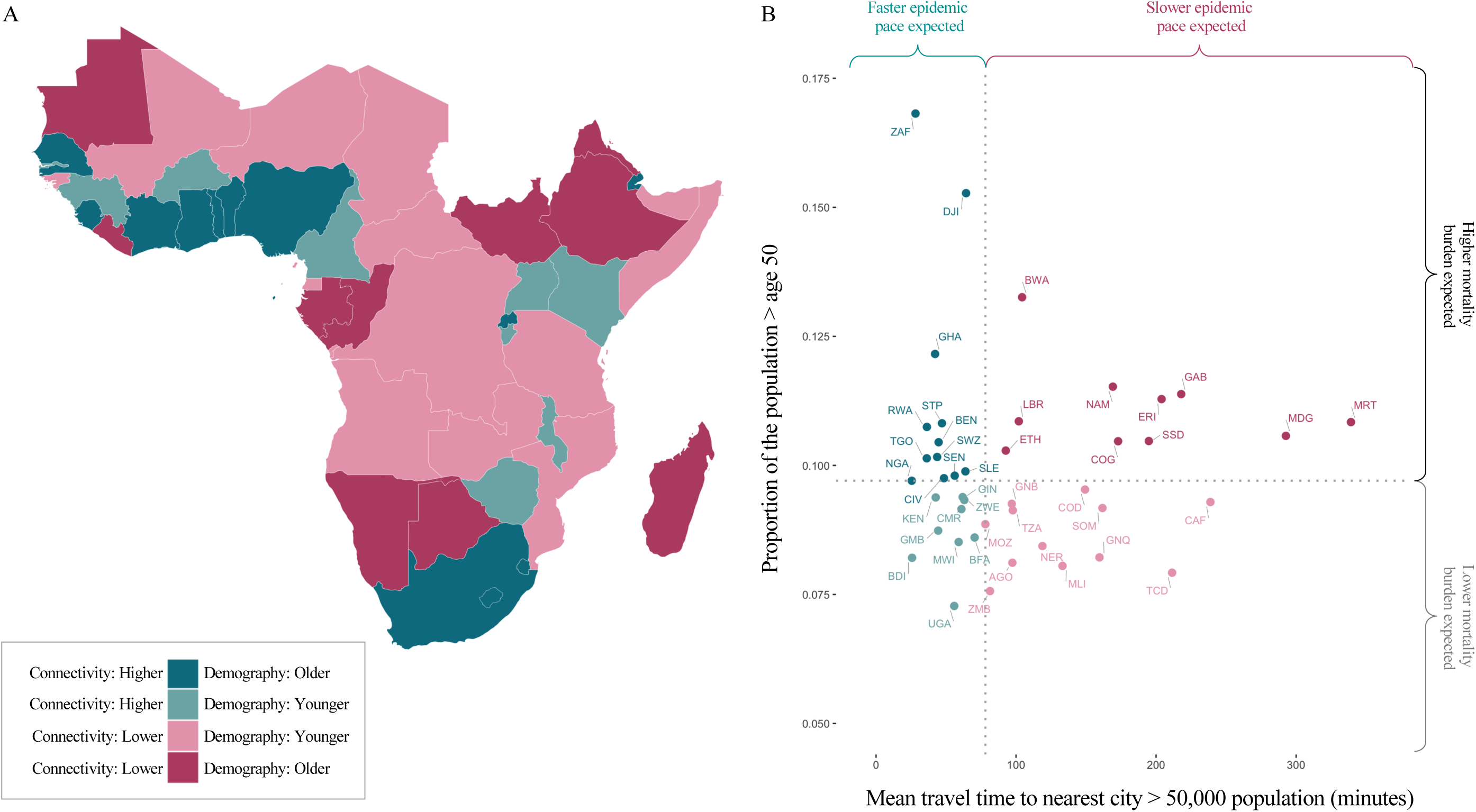
Expected pace versus expected burden at the national level in SARS-CoV-2 outbreaks in sub-Saharan Africa. Countries are colored by with respect to indicators of their expected epidemic pace (using as an example subnational connectivity in terms of travel time to nearest city) and potential burden (using as an example the proportion of the population over age 50). **A:** In pink, countries with less connectivity (i.e., less synchronous outbreaks) relative to the median among SSA countries; in blue, countries with more connectivity; darker colors show countries with older populations (i.e., a greater proportion in higher risk age groups). **B**: Dotted lines show the median; in the upper right, in dark pink, countries are highlighted due to their increased potential risk for an outbreak to be prolonged (see metapopulation model methods) and high burden (see burden estimation methods).

## Online Content

Methods and additional figures are available in the supplementary materials. In addition, high resolution maps and further visualizations of the risk indicators and simulations studied here can be accessed online through an interactive tool: Link to SSA-SARS-CoV-2 online companion tool: https://labmetcalf.shinyapps.io/covid19-burden-africa/

## Data Availability

Data and code have been deposited into a publicly available GitHub repository: https://github.com/labmetcalf/SSA-SARS-CoV-2
High resolution maps and further visualizations of the risk indicators and simulations studied here can be accessed online through an interactive tool:
https://labmetcalf.shinyapps.io/covid19-burden-africa/

## Data Availability

All data have been deposited into a publicly available GitHub repository: Link to GitHub repository containing data and code: https://github.com/labmetcalf/SSA-SARS-CoV-2

## Code Availability

All code has been deposited into the publicly available GitHub repository (same as above): Link to GitHub repository containing data and code: https://github.com/labmetcalf/SSA-SARS-CoV-2

## Acknowledgements

REB is supported by the Cooperative Institute for Modeling the Earth System (CIMES). AA acknowledges support from the NIH Medical Scientist Training Program 1T32GM136577. AJT is funded by the BMGF (OPP1182425, OPP1134076 and INV-002697). MB is funded by NWO Rubicon grant 019.192EN.017.

## Author contributions

*Conceptualization*: BLR, AA, REB, MB, WWD, KM, IFM, NVM, AR, MR, JR, TR, FR, WY, BTG, CJT, CJEM; *Data curation*: BLR, MR, MB, WWD, WY; *Formal analysis*: BLR, AA, MB, MR, REB; *Methodology*: BLR, MR, MB, REB, CJEM, BTG; *Software and Shiny app online tool*: BLR, MR, MB, REB, WY; *Visualization*: BLR, MR, MB, REB, WY; *Writing* – original draft: BLR, CJEM; *Writing – reviewing and editing*: BLR, AA, REB, MB, WWD, KM, IFM, NVM, AR, MR, JR, TR, FR, WY, BTG, CJT, CJEM

## Additional Information

Supplementary Information is available for this paper. Correspondence and requests for materials should be addressed to BLR

## Data and materials availability

All materials are available in the online content

## Competing interests

The authors declare no competing interests

Prevention Access to care Human mobility and travel

## Supplementary Materials Outline

<u>A1 Reported SARS-CoV-2 case counts, mortality, and testing in sub-Saharan Africa as of June 2020</u>

Table S1: Sub-Saharan Africa country codes, case counts, and testing Figure S1: Variation between SSA countries in testing and reporting rates

<u>A2 Synthesizing factors hypothesized to increase or decrease SARS-CoV-2 epidemic risk in SSA</u>

Table S2: Dimensions of risk and expected direction of effect on SARS-CoV-2 transmission or burden in sub-Saharan Africa (SSA) relative to higher latitude countries

Table S3: Variables and data sources

Figure S2: Year of most recent data available for variables compared between global regions

Figure S3: Variation among sub-Saharan African countries in determinants of SARS-CoV-2 risk by variable (a subset of variables is shown in Figure 2 in the main text)

Figure S4: Variation among sub-Saharan African countries in determinants of SARS-CoV-2 mortality risk by category (subsets of variables are shown in Figure 3 in the main text)

Data File 1: Data for all compiled indicators

<u>A3 Principal component analysis (PCA) of variables considered</u>

Figure S5: PCAs of all variables and category specific subsets of variables Data File 2: GDP, GINI Index, and tests completed data for PCA visualizations

<u>A4 Evaluating the burden emerging from the severity of infection outcome</u> Table S4: Sources of age-stratified infection fatality ratio (*IFR*) estimates Figure S6: Age profiles of comorbidities in sub-Saharan Africa countries

<u>A5 International air travel to sub-Saharan Africa</u>

Table S5: Arrivals to SSA airports by the number of passenger seats and status of the SARS-CoV-2 pandemic at the origin at the time of travel

<u>A6 Subnational connectivity among countries in sub-Saharan Africa</u>

Metapopulation model methods Figure S7: Pace of the outbreak

Figure S8: Cases and testing vs. the pace of the outbreak

<u>A7 Modeling epidemic trajectories in scenarios where transmission rate depends on climate</u>

Data on climate variation in SSA Climate model methods

## A1 Reported SARS-CoV-2 case counts, mortality, and testing in sub-Saharan Africa as of June 2020

### 1.1. Variables and data sources for testing data

The numbers of reported cases, deaths, and tests for the 48 studied sub-Saharan Africa (SSA) countries (**Table S1**) were sourced from the Africa Centers for Disease Control (CDC) dashboard on June 30, 2020 (https://africacdc.org/covid-19/). Africa CDC obtains data from the official Africa CDC Regional Collaborating Centre and member state reports. Differences in the timing of reporting by member states results in some variation in recency of data within the centralized Africa CDC repository, but the data should broadly reflect the relative scale of testing and reporting efforts across countries.

The countries or member states within SSA in this study follow the United Nations and Africa CDC listed regions of Southern, Western, Central, and Eastern Africa (not including Sudan). From the Northern Africa region, Mauritania is included in SSA.

For comparison to non-SSA countries, the number of reported cases in other geographic regions were obtained from the Johns Hopkins University Coronavirus Resource Center on June 30, 2020 (https://coronavirus.jhu.edu/map.html).

Case fatality ratios (*CFR*s) were calculated by dividing the number of reported deaths by the number of reported cases and expressed as a percentage. Positivity was calculated by dividing the number of reported cases by the number of reported tests. Testing and case rates were calculated per 100,000 population using population size estimates for 2020 from the United Nations Population Division ^38^. As reported confirmed cases are likely to be a significant underestimate of the true number of infections, *CFR*s may be a poor proxy for the infection fatality ratio (*IFR*), defined as the proportion of infections that result in mortality ^4^.

### 1.2 Variation in testing and mortality rates

Testing rates among SSA countries varied by multiple orders of magnitude: the number of tests completed per 100,000 population ranged from 6.50 in Tanzania to 13,508.13 in Mauritius (**Figure S1A**). The number of reported infections (i.e., positive tests) was strongly correlated with the number of tests completed (Pearson’s correlation coefficient, *r* = 0.9667, *p* < 0.001) (**Figure S1B**). As of June 30, 2020, no deaths due to SARS-CoV-2 were reported to the Africa CDC for five SSA countries (Eritrea, Lesotho, Namibia, Seychelles, Uganda). Among countries with at least one reported death, *CFR* varied from 0.22% in Rwanda to 8.54% in Chad (**Figure S1C**). Limitations in the ascertainment of infection rates and the rarity of reported deaths (e.g., median number of reported deaths per SSA country was 25.5), indicate that the data are insufficient to determine country specific *IFR*s and *IFR* by age profiles. As a result, global *IFR* by age estimates were used for the subsequent analyses in this study.

**Table S1.** Sub-Saharan Africa country country codes, case counts, and testing as of June 30, 2020

| Country Name | Country Code | Cases<sup>a</sup> | Deaths<sup>a</sup> | Tests<sup>a</sup> | Population<sup>b</sup> | Cases per 100k<sup>c</sup> | Tests per 100k<sup>c</sup> | Positivity (%) | CFR (%) |
| --- | --- | --- | --- | --- | --- | --- | --- | --- | --- |
| Angola | AGO | 267 | 11 | 22895 | 32866268 | 0.81 | 69.66 | 1.17 | 4.12 |
| Benin | BEN | 1187 | 19 | 20014 | 12123198 | 9.79 | 165.09 | 5.93 | 1.60 |
| Botswana | BWA | 89 | 1 | 36868 | 2351625 | 3.78 | 1567.77 | 0.24 | 1.12 |
| Burkina Faso | BFA | 959 | 53 | 9040 | 20903278 | 4.59 | 43.25 | 10.61 | 5.53 |
| Burundi | BDI | 170 | 1 | 2359 | 11890781 | 1.43 | 19.84 | 7.21 | 0.59 |
| Cameroon | CMR | 12592 | 313 | 80000 | 26545864 | 47.43 | 301.37 | 15.74 | 2.49 |
| Cabo Verde | CPV | 1165 | 12 | 22665 | 555988 | 209.54 | 4076.53 | 5.14 | 1.03 |
| Central Africa Republic | CAF | 3429 | 45 | 23208 | 4829764 | 71.00 | 480.52 | 14.78 | 1.31 |
| Chad | TCD | 866 | 74 | 4633 | 16425859 | 5.27 | 28.21 | 18.69 | 8.55 |
| Comoros | COM | 293 | 7 | 1173 | 869595 | 33.69 | 134.89 | 24.98 | 2.39 |
| Côte d'Ivoire | CIV | 9101 | 66 | 48340 | 26378275 | 34.50 | 183.26 | 18.83 | 0.73 |
| Congo (DRC) | COD | 6939 | 167 | 24657 | 89561404 | 7.75 | 27.53 | 28.14 | 2.41 |
| Djibouti | DJI | 4656 | 53 | 46108 | 988002 | 471.25 | 4666.79 | 10.10 | 1.14 |
| Equatorial Guinea | GNQ | 2001 | 32 | 16000 | 1402985 | 142.62 | 1140.43 | 12.51 | 1.60 |
| Eritrea | ERI | 191 | 0 | 7943 | 3546427 | 5.39 | 223.97 | 2.40 | 0.00 |
| Eswatini | SWZ | 781 | 11 | 11094 | 1160164 | 67.32 | 956.24 | 7.04 | 1.41 |
| Ethiopia | ETH | 5846 | 103 | 250604 | 114963583 | 5.09 | 217.99 | 2.33 | 1.76 |
| Gabon | GAB | 5209 | 40 | 34774 | 2225728 | 234.04 | 1562.37 | 14.98 | 0.77 |
| Gambia | GMB | 45 | 2 | 2947 | 2416664 | 1.86 | 121.94 | 1.53 | 4.44 |
| Ghana | GHA | 17351 | 112 | 294867 | 31072945 | 55.84 | 948.95 | 5.88 | 0.65 |
| Guinea | GIN | 5291 | 30 | 33737 | 13132792 | 40.29 | 256.89 | 15.68 | 0.57 |
| Guinea Bissau | GNB | 1614 | 21 | 8056 | 1967998 | 82.01 | 409.35 | 20.03 | 1.30 |
| Kenya | KEN | 6190 | 144 | 167417 | 53771300 | 11.51 | 311.35 | 3.70 | 2.33 |
| Lesotho | LSO | 27 | 0 | 3000 | 2142252 | 1.26 | 140.04 | 0.90 | 0.00 |
| Liberia | LBR | 768 | 34 | 6125 | 5057677 | 15.18 | 121.10 | 12.54 | 4.43 |

412 (Table S1 continued)
| Country Name | Country Code | Cases<sup>a</sup> | Deaths<sup>a</sup> | Tests<sup>a</sup> | Population<sup>b</sup> | Cases per 100k<sup>c</sup> | Tests per 100k<sup>c</sup> | Positivity (%) | CFR (%) |
| --- | --- | --- | --- | --- | --- | --- | --- | --- | --- |
| Madagascar | MDG | 2138 | 20 | 21444 | 27691019 | 7.72 | 77.44 | 9.97 | 0.94 |
| Malawi | MWI | 1152 | 13 | 13369 | 19129955 | 6.02 | 69.89 | 8.62 | 1.13 |
| Mali | MLI | 2147 | 113 | 12869 | 20250834 | 10.60 | 63.55 | 16.68 | 5.26 |
| Mauritania | MRT | 4149 | 126 | 39398 | 4649660 | 89.23 | 847.33 | 10.53 | 3.04 |
| Mauritius | MUS | 341 | 10 | 171792 | 1271767 | 26.81 | 13508.13 | 0.20 | 2.93 |
| Mozambique | MOZ | 859 | 5 | 28586 | 31255435 | 2.75 | 91.46 | 3.00 | 0.58 |
| Namibia | NAM | 183 | 0 | 8706 | 2540916 | 7.20 | 342.63 | 2.10 | 0.00 |
| Niger | NER | 1074 | 67 | 6555 | 24206636 | 4.44 | 27.08 | 16.38 | 6.24 |
| Nigeria | NGA | 24567 | 565 | 130164 | 206139587 | 11.92 | 63.14 | 18.87 | 2.30 |
| Congo (ROC) | COG | 1245 | 40 | 11790 | 5518092 | 22.56 | 213.66 | 10.56 | 3.21 |
| Rwanda | RWA | 900 | 2 | 137751 | 12952209 | 6.95 | 1063.53 | 0.65 | 0.22 |
| São Tomé and Príncipe | STP | 713 | 13 | 17773 | 219161 | 325.33 | 8109.56 | 4.01 | 1.82 |
| Senegal | SEN | 6698 | 108 | 76343 | 16743930 | 40.00 | 455.94 | 8.77 | 1.61 |
| Seychelles | SYC | 77 | 0 | 704 | 98340 | 78.30 | 715.88 | 10.94 | 0.00 |
| Sierra Leone | SLE | 1427 | 60 | 9973 | 7976985 | 17.89 | 125.02 | 14.31 | 4.20 |
| Somalia | SOM | 2894 | 90 | 11807 | 15893219 | 18.21 | 74.29 | 24.51 | 3.11 |
| South Africa | ZAF | 138134 | 2456 | 1567084 | 59308690 | 232.91 | 2642.25 | 8.81 | 1.78 |
| South Sudan | SSD | 2006 | 37 | 10630 | 11193729 | 17.92 | 94.96 | 18.87 | 1.84 |
| Tanzania | TZA | 509 | 21 | 3880 | 59734213 | 0.85 | 6.50 | 13.12 | 4.13 |
| Togo | TGO | 642 | 14 | 30316 | 8278737 | 7.75 | 366.19 | 2.12 | 2.18 |
| Uganda | UGA | 870 | 0 | 186200 | 45741000 | 1.90 | 407.07 | 0.47 | 0.00 |
| Zambia | ZMB | 1531 | 21 | 53370 | 18383956 | 8.33 | 290.31 | 2.87 | 1.37 |
| Zimbabwe | ZWE | 567 | 6 | 66712 | 14862927 | 3.81 | 448.85 | 0.85 | 1.06 |

<sup>a</sup> Data from Africa CDC as of June 30, 2020 (https://africacdc.org/covid-19/)
<sup>b</sup> Data from UN Population Division UNPOP (2019 revision) estimates of population by single calendar year (2020)<sup>38</sup>
<sup>c</sup> Rates per 100,000 population

**Figure S1.**
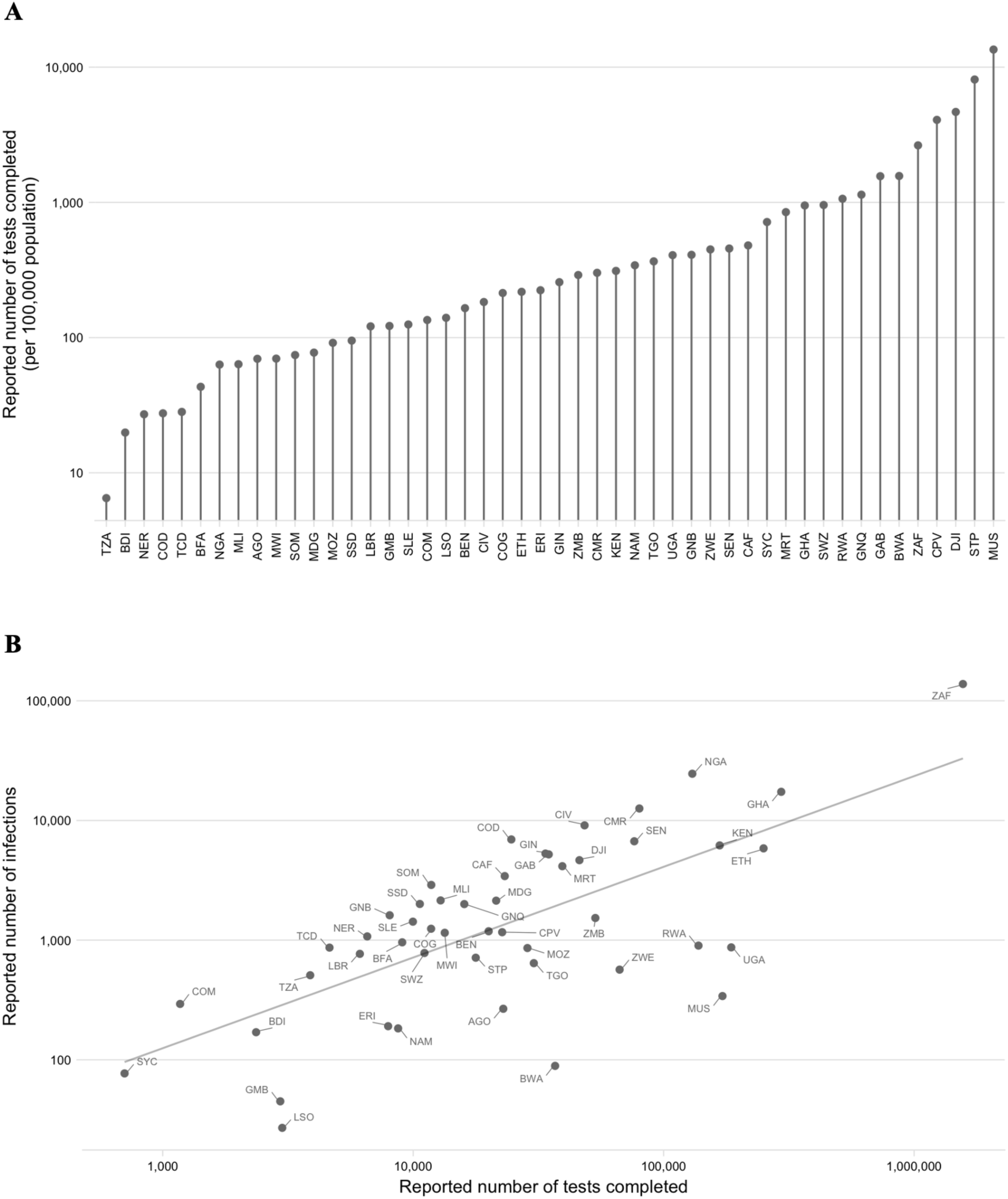
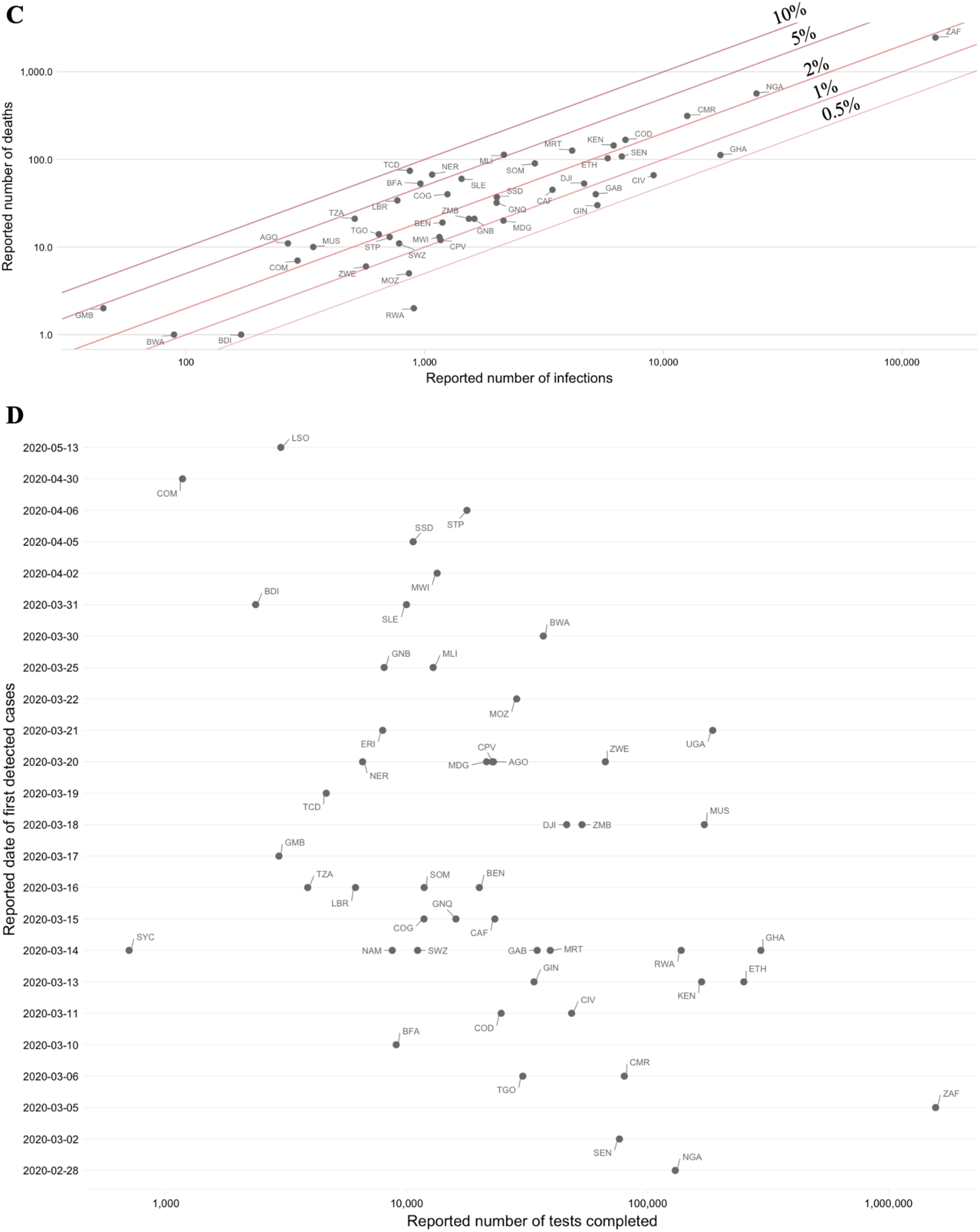
Variation between SSA countries in testing and reporting rates as of June 30, 2020. **A:** Reported number of tests completed per country as of June 2020 (source: Africa CDC). **B**: Number of infections (*I*) per reported number of tests (*T*); line shows linear regression: *I* = 8.454×10^−2^×*T* − 8.137×10^2^ (*R*^*2*^ = 0.933, *p* < 0.001). **C**: Reported infections and deaths for sub-Saharan African countries with case fatality ratios (*CFR*s) shown as diagonal lines. **D**: Date of first detection per number of reported tests

## A2 Methods: Synthesizing factors hypothesized to increase or decrease SARS-CoV-2 epidemic risk in SSA

### 2.1. Variable selection and data sources for variables hypothesized to associate with an increased probability of severe clinical outcomes for an infection

To characterize epidemic risk, defined as potential SARS-CoV-2 related morbidity and mortality, we first synthesized factors hypothesized to influence risk in SSA settings (**Table S2**). Early during the pandemic, evidence suggested that age was an important risk factor associated with morbidity and mortality associated with SARS-CoV-2 infection ^39^, a pattern subsequently confirmed across settings ^2,9,40^. Associations between SARS-CoV-2 mortality and comorbidities including hypertension, diabetes, and cardiovascular disease emerged early ^39^; and have been observed across settings, with further growing evidence for associations with obesity ^9,41^, severe asthma ^9^, and respiratory effects of pollution ^42^.

Many possible sources of bias complicate interpretation of these associations ^43^, and while they provide a useful baseline, inference is also likely to change as the pandemic advances. To reflect this, our analysis combines a number of high level variables likely to broadly encompass these putative risk factors (e.g., non-communicable disease (NCD) related mortality and health life expectancy) with more specific measures encompassed in evidence to date (e.g., prevalence of diabetes, obesity, and respiratory illness such as Chronic Obstructive Pulmonary Disease (COPD)). We also include measures relating to infectious diseases, undernourishment, and anemia given their interaction and effects in determining health status in these settings ^44^.

Data on the identified indicators were sourced in May 2020 from the World Health Organization (WHO) Global Health Observatory (GHO) database (https://www.who.int/data/gho), World Bank (https://data.worldbank.org/), and other sources detailed in **Table S3**. National level demographic data (population size and age structure) was sourced from United Nations World Population Prospects (UNPOP) ^38^ and data on subnational variation in demography was sourced from WorldPop ^25^. Household size data was defined by the mean number of individuals in a household with at least one person aged > 50 years, taken from the most recently available demographic health survey (DHS) data ^45^. All country level data for all indicators can be found online at the **SSA-SARS-CoV-2-tool** (https://labmetcalf.shinyapps.io/covid19-burden-africa/).

Comparisons of national level estimates sourced from WHO and other sources are affected by variation within countries and variation in the uncertainty around estimates from different geographical areas. To assess potential differences in data quality between geographic areas we compared the year of most recent data for variables (**Figure S2**). The mean (range varied from 2014.624 to 2014.928 by region) and median year (2016 for all regions) of the most recent data varied little between regions. To account for uncertainty associated in the estimates available for a single variable, we also include multiple variables per category (e.g., demographic and socio-economic factors, comorbidities, access to care) to avoid reliance on a single metric. This allows exploring variation between countries across a broad suite of variables likely to be indicative of the different dimensions of risk.

Although including multiple variables that are likely to be correlated (see PCA methods below for further discussion) would bias inference of cumulative risk in a statistical framework, we do not attempt to quantitatively combine risk across variables for a country, nor project risk based on the variables included here. Rather, we characterize the magnitude of variation among countries for these variables (see **Figure 2** in the main text for a subset of the variables; **Figure 3B** for bivariate risk maps following ^46^) and then explore the range of outcomes that would be expected under scenarios where *IFR* increases with age at different rates (see **Figure 3** in the main text).

### 2.2. Variable selection and data sources for variables hypothesized to modulate the rate of viral spread

In addition to characterizing variation among factors likely to modulate burden, we also synthesize data sources relevant to the rate of viral spread, or pace, for the SARS-CoV-2 pandemic in SSA. Factors hypothesized to modulate viral transmission and geographic spread include climatic factors (e.g., specific humidity), access to prevention measures (e.g., handwashing), and human mobility (e.g., international and domestic travel). **Table S2** outlines the dimensions of risk selected and references the previous studies relevant to the selection of these factors.

Climate data was sourced from the global, gridded ERA5 dataset ^47^ where model data is combined with global observation data (see **Section A7** for details).

International flight data was obtained from a custom report from OAG Aviation Worldwide (UK) and included the departure location, airport of arrival, date of travel, and number of passenger seats for flights arriving to 113 international airports in SSA (see **Section A5**).

As an estimate of connectivity within subregions of countries, the population weighted mean travel time to the nearest city with a population greater than 50,000 was determined; details are provided in **Section A6**. To obtain a set of measures that broadly represent connectivity within different countries in the region, friction surfaces from ref^24^ were used to obtain estimates of the connectivity between different administrative level 2 units within each country. Details of this, alongside the metapopulation model framework used to simulate viral spread with variation in connectivity are in **Section A6**.

**Figure 2** in the main text shows variation among SSA countries for four of the variables; **Figure S3** shows variation for all variables. **Figure 3** in the main text shows variation for a subset of the comorbidity and access to care indicators as a heatmap; **Figure S4** shows variation for all the variables (both also available online at the **SSA-SARS-CoV-2-tool** (https://labmetcalf.shinyapps.io/covid19-burden-africa/)).

**Table S2.** Hypothesized dimensions of risk and expected direction of effect on SARS-CoV-2 transmission or burden in sub-Saharan Africa (SSA) relative to higher latitude countries

| Dimension of risk | Factors hypothesized to **decrease** transmission or burden in sub-Saharan Africa relative to other geographic areas | Factors hypothesized to **increase** transmission or burden in sub-Saharan Africa relative to other geographic areas |
| --- | --- | --- |
| *(A) Demographic and socio-economic characteristics in SSA* | Younger populations, and thus a smaller proportion of individuals in the older age groups that experience the highest mortality<sup>2-4</sup> | A larger proportion of urban populations living in dense settings, which may result in higher transmission<sup>48</sup>; higher contact with older individuals as a result of multi-generation households<sup>14</sup> |
| *(B) Comorbidities in SSA* | Lower rates of some comorbidities that have been associated with risk of worse outcomes, e.g., obesity<sup>9,41</sup> | Higher rates of NCDs such as hypertension or COPD<sup>39</sup>, which are associated with worse outcomes; and a potential role for as yet undescribed interactions e.g., with anemia, or high prevalence infectious diseases |
| *(C) Climate in SSA* | Warmer, wetter climates on average driving reduced transmission<sup>1,49</sup> |  |
| *(D) Capacity to deploy prevention measures in SSA* | Experience with previous outbreak response which may yield more rapid and nimble approaches to reducing transmission<sup>21,22</sup> | Lower access to handwashing<sup>50,51</sup> and other prevention options such as self-isolation<sup>52</sup>, increasing transmission<br>Subregions of countries with reduced governance infrastructure<sup>53</sup> |
| *(E) Access to healthcare in SSA* |  | Larger variation in access to and coverage of health systems<sup>54</sup> including fewer medical staff and facilities such as hospital beds<sup>14</sup> increasing burden<br>Increased vulnerability to disruption of routine health services (e.g.,<sup>31</sup>)<br>Limited testing capacity<sup>55</sup> reducing the capacity to identify and interrupt chains of transmission |
| *(F) Human mobility and travel in SSA* | Fewer viral importations due to reduced frequency of international travel<sup>29,30</sup><br>Decreased rate of internal spread due to less connectivity within countries<sup>56</sup> |  |

**Table S3.** Variables and data sources for indicators of SARS-CoV-2 epidemic risk in sub-Saharan Africa

| ID | Variable | Source | Hypothesized association(s) with SARS-CoV-2 outcomes |
| --- | --- | --- | --- |
| (A) Demographic and socio-economic characteristics |  |  |  |
| A1 | Human population size; Proportion of population over age 50 (%) from the UN Population Division UNPOP (2019 revision) estimates of population by single calendar year (2020), age, and country | UN<sup>38</sup> | Morbidity and mortality observed to increase with age (e.g., <sup>2-4</sup>) |
| A2 | Subnational spatial variation in the distribution of the human population and age structure | WorldPop |  |
| A3 | Household size: Mean household size for households with an individual over age 50 | DHS | Proxy for social contact rate for the elderly population at higher risk for SARS-CoV-2 morbidity and mortality <sup>14</sup> |
| A4 | Proportion of households with an individual over age 50 | DHS |  |
| A5 | Health life expectancy (HALE) at age 60 (years) | WHO | Proxy for baseline health status of elderly population |
| A6 | Proportion of population below the poverty line (%) | World Bank | More severe clinical outcomes associated with poverty; A proxy for access to advanced care <sup>57,58</sup> |
| A7 | Proportion of the urban population living in crowded, low quality housing (defined as households lacking one or more of the following conditions: access to improved water, access to improved sanitation, sufficient living area, and durability of housing) (%) | World Bank | Indicator of capacity for prevention (e.g., through handwashing); Transmission observed to increase with crowding <sup>48</sup> |
| A8 | Gross domestic product (GDP) per capita | World Bank | Used in PCA analysis (see below) as an indicator of socio-economic status at the national level |
| A9 | GINI index, a measure of inequality in the distribution of income | World Bank |  |
| (B) Comorbidities: General and nutrition related non-communicable diseases (NCDs) |  |  |  |
| B1 | NCDs overall mortality per 100 000 popn, age-standardized | WHO | Indicator of NCD burden in population; Comorbidities increase probability of severe clinical outcomes |
| B2 | Cardiovascular disease related mortality per age group (annual deaths attributable per 100,000 population) | GDB 2017<sup>59</sup> |  |
| B3 | Diabetes prevalence among ages 20-79 (%) | World Bank | Increases probability of severe clinical outcomes |
| B4 | Diabetes related mortality per age group (annual deaths attributable per 100,000 population) | GDB 2017<sup>59</sup> |  |

(Table S3 continued)
| ID | Variable | Source | Hypothesized association(s) with SARS-CoV-2 outcomes |
| --- | --- | --- | --- |
| (B) Comorbidities: General and nutrition related non-communicable diseases (NCDs) |  |  |  |
| B5 | Raised glucose prevalence, age-standardized (%) | WHO | Indicator of metabolic disease risk; Metabolic disease increases probability of severe clinical outcomes |
| B6 | Raised blood pressure prevalence, age-standardized (%) | WHO |  |
| B7 | Raised cholesterol prevalence, age-standardized (%) | WHO |  |
| B8 | Overweight prevalence among adults, age-standardized (%) | WHO |  |
| B9 | Anemia prevalence among non-pregnant women (%) | WHO | Indicator of poor nutritional status; Poor nutritional status may increase probability of severe clinical outcomes |
| B10 | Undernourishment prevalence (%) | WHO |  |
| (B) Comorbidities: NCDs related to respiratory system and pollution |  |  |  |
| B11 | Annual mean PM2.5 exposure in urban areas (ug/m3) | WHO | Exposure to air pollution increases mortality <sup>42</sup> |
| B12 | Lung, tracheal, and esophageal cancer mortality per 100 000 popn, age-standardized | WHO | Indicator of prevalence and management of chronic disease and inflammation affecting the respiratory tract |
| B13 | Chronic respiratory diseases (excluding asthma) related mortality per age group (annual deaths attributable per 100,000 population) | GDB 2017<sup>59</sup> |  |
| B14 | COPD mortality per 100 000 popn, age-standardized | WHO |  |
| (B) Comorbidities: Infectious diseases |  |  |  |
| B15 | Respiratory infections mortality per 100 000 popn, age-standardized | WHO | Indicator of prevalence and management of infectious disease affecting the respiratory tract |
| B16 | TB incidence per 100 000 popn | World Bank | Indicator of susceptibility to respiratory infections and immune suppression |
| B17 | HIV prevalence among ages 15-49 (%) | World Bank | Indicator of immunosuppressed population |
| (C) Climate |  |  |  |
| C1 | Seasonal change in specific humidity (in selected urban centers) | ERA5<sup>47</sup> | Transmission rate of coronaviruses may decline with humidity |
| (D) Capacity to deploy prevention measures |  |  |  |
| D1 | Proportion of urban popn with basic handwashing facilities with water and soap at home (%) | WHO | Handwashing observed to reduce infection rates for respiratory pathogens |
| D2 | Proportion of the population with access to a handwashing station with soap and water in 2019 | Ref<sup>60</sup> |  |

(Table S3 continued)
| ID | Variable | Source | Hypothesized association(s) with SARS-CoV-2 outcomes |
| --- | --- | --- | --- |
| (D) Capacity to deploy prevention measures |  |  |  |
| D3 | Proportion of 1 year olds receiving full immunization coverage (%) | WHO | Proxy for coverage of routine health services |
| D4 | Reported number of completed tests reported for SARS-CoV-2 infection as of June 30, 2020 | Africa CDC | Indicator of surveillance capacity |
| (E) Access to healthcare in SSA |  |  |  |
| E1 | Proportion of children with pneumonia symptoms taken to a health facility (%) | WHO | Proxy for access to medical care and care seeking |
| E2 | Subnational spatial variation in the probability of seeking treatment for fever at public facilities | Ref<sup>61</sup> |  |
| E3 | Proportion of births attended by skilled staff (%) | World Bank |  |
| E4 | Nurses and midwives per 100 000 popn | World Bank | Indicators of treatment capacity |
| E5 | Physicians per 100 000 popn | World Bank |  |
| E6 | Hospitals per 100 000 popn | World Bank |  |
| E7 | Hospital beds 100 000 popn | World Bank |  |
| E8 | Health expenditure per capita in (USD) | WHO | Proxy for health system resources; A significant predictor of intensive care unit (ICU) capacity <sup>62</sup> |
| E9 | Proportion of health expenditures that are out-of-pocket (%) | WHO |  |
| (F) Human mobility and travel: International |  |  |  |
| F1 | Estimated number of international passengers arriving at SSA airports from January-April 2020 | OAG | Indicator of the timing and number of introductions of SARS-CoV-2 |
| F2 | Estimated number of international passengers arriving at SSA airports from January-April 2020 by SARS-CoV-2 status at departure location | OAG |  |

(Table S3 continued)
| ID | Variable | Source | Hypothesized association(s) with SARS-CoV-2 outcomes |
| --- | --- | --- | --- |
| *(F) Human mobility and travel: Domestic* |  |  |  |
| F3 | National population-weighted mean travel time to the nearest city (national mean of indicator F4) | Ref<sup>63</sup> | Indicator of connectivity within countries; A proxy for the rate of human mobility |
| F4 | Population-weighted mean travel time to the nearest city (population > 50,000) for administrative level 2 units | Ref<sup>63</sup> |  |
| F5 | Relative costs of travel between centroids of administrative level 2 derived from friction surfaces obtained by integrating data on travel infrastructure (Open Street Map, land cover types, etc). | Ref<sup>24</sup> |  |

**Figure S2.**
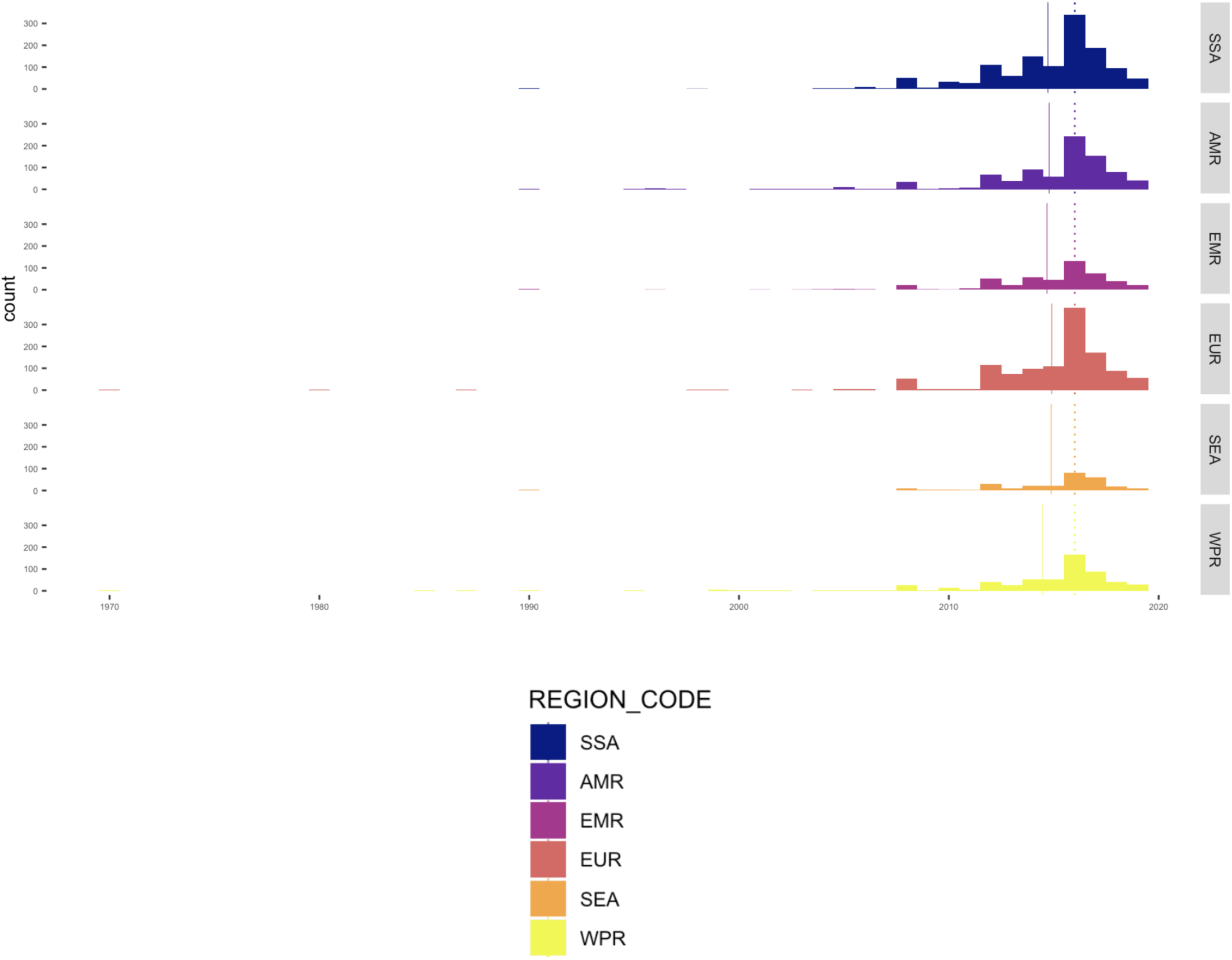
Year of most recent data available for variables compared between global regions. Dotted vertical line shows regional median; solid vertical line shows regional mean. Note that most data comes from 2015-2019 (median = 2016, mean = 2014.62-2014.93).

**Figure S3. Variation among sub-Saharan African countries in determinants of SARS-CoV-2 risk by variable**

A subset of variables is shown in Figure 2A-D in the main text, the remaining variables are shown in supplementary file “Figure S3 compiled.pdf” and available online: **SSA-SARS-CoV-2-tool** (https://labmetcalf.shinyapps.io/covid19-burden-africa/)

**Figure S4.**
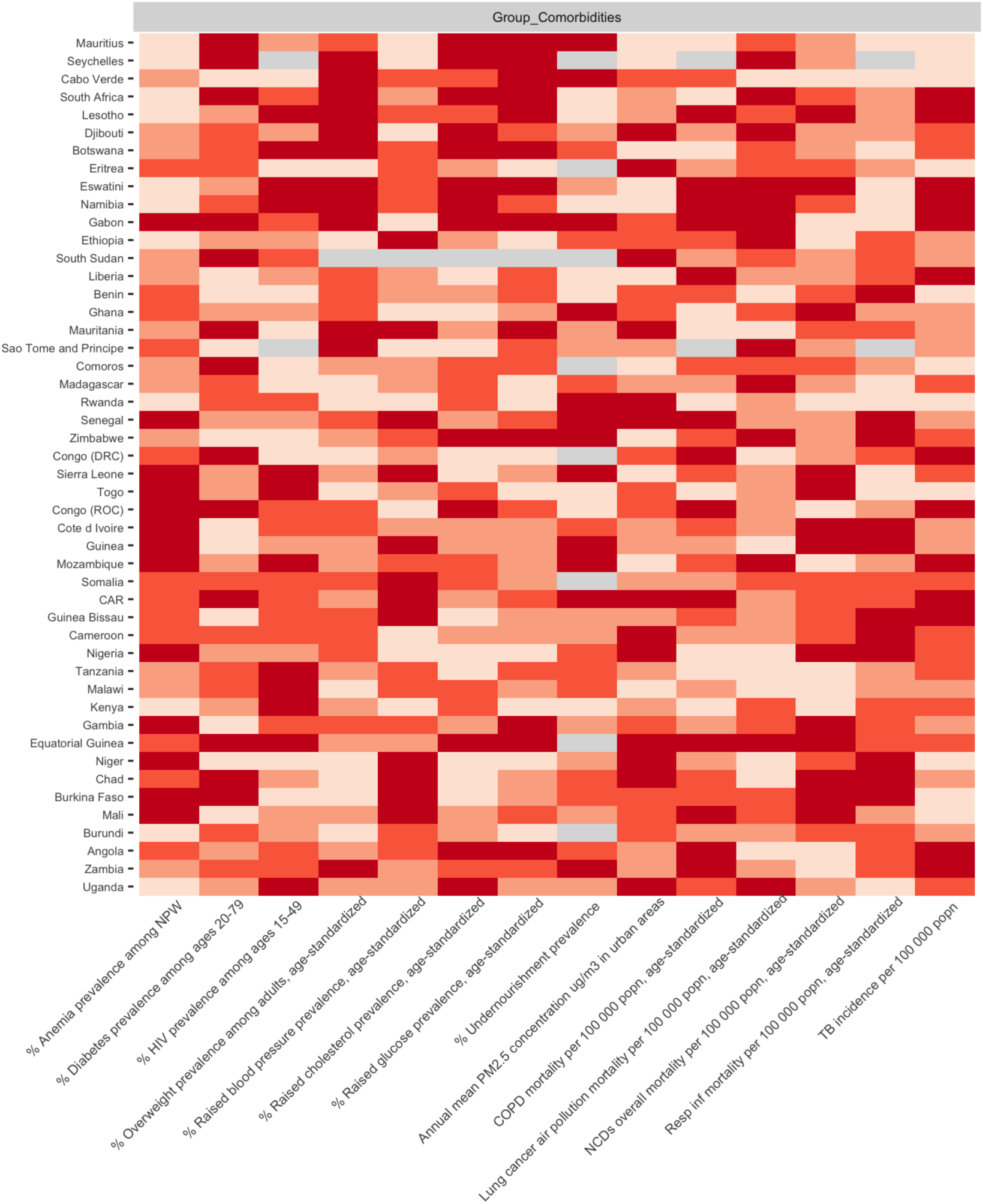
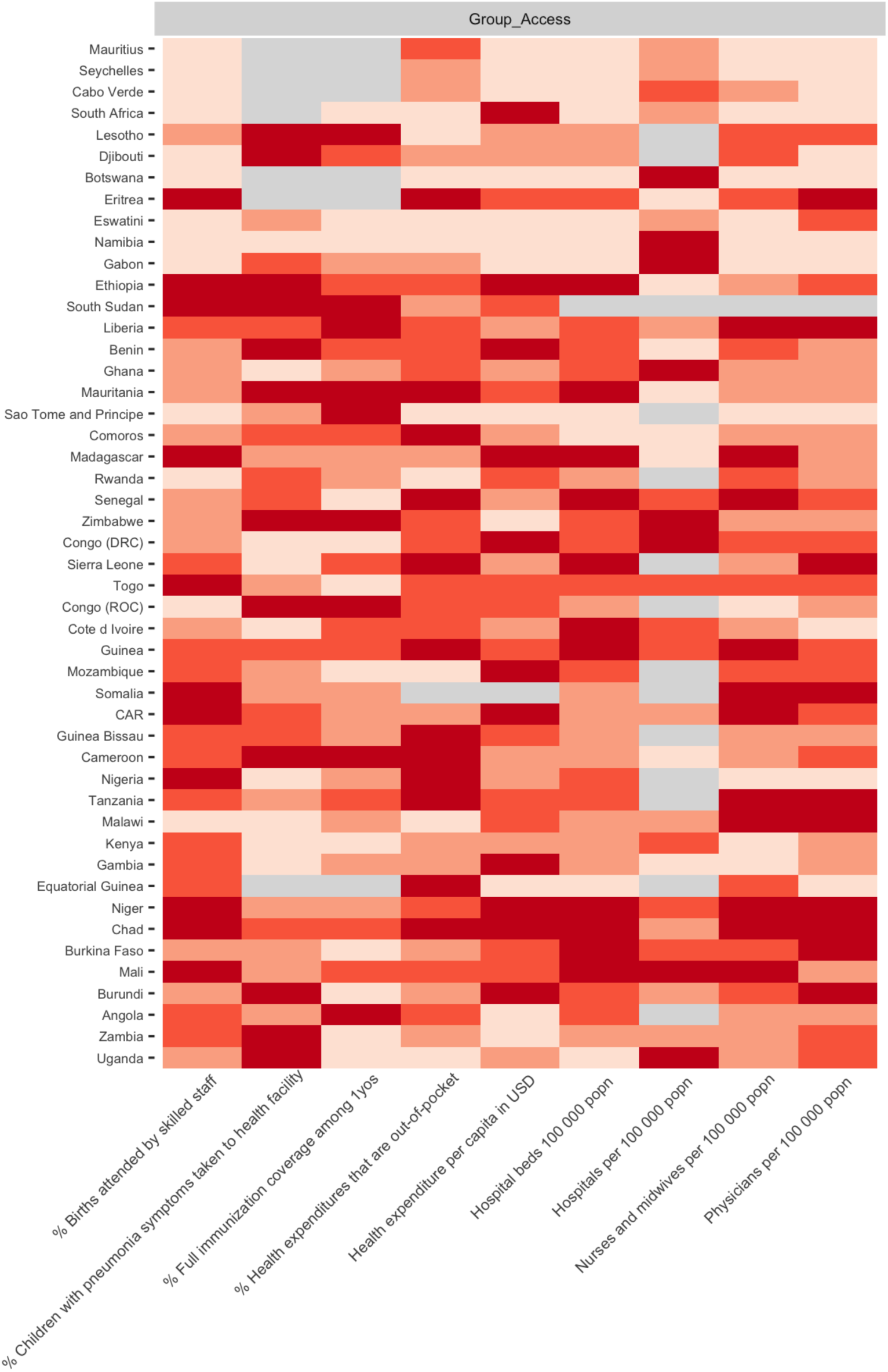
Variation among sub-Saharan African countries in determinants of SARS-CoV-2 mortality risk by category. A subset of variables is shown in Figure 3D-E in the main text, the remaining variables are shown and available online: **SSA-SARS-CoV-2-tool** (https://labmetcalf.shinyapps.io/covid19-burden-africa/) **A**: Select national level indicators; estimates of increased comorbidity burden (e.g., higher prevalence of raised blood pressure) shown with darker red for higher risk quartiles Countries missing data for an indicator (NA) are shown in gray. For comparison between countries, estimates are age-standardized where applicable (see **Table S3** for details) **B**: Select national level indicators; estimates of reduced access to care (e.g., fewer hospitals) shown with darker red for higher risk quartiles Countries missing data for an indicator (NA) are shown in gray. For comparison between countries, estimates are age-standardized where applicable (see **Table S3** for details)

## A3 Principal component analysis (PCA) of variables considered

### 3.1. Selection of data and variables

The 29 national level variables from **Table S3** were selected for principal component analysis (PCA). We conducted further PCA on the subset of eight indicators related to access to healthcare (Category E) and the 14 national indicators variables related to comorbidities (Category B).

We excluded disaggregated sub-national spatial variation data (variables A2, C1, E2, and Category F), disaggregated or redundant variables derived from already included variables (variables A4 and D2), and disaggregated age-specific disease data from IHME global burden of disease study (variables B2, B4, and B13) from PCA analysis. COVID-19 tests per 100,000 population (variable D4, **Table S1**), per capita gross domestic product (GDP) (Variable A8), and the GINI index of wealth inequality (Variable A9) were used to visualize patterns among sub-Saharan Africa countries.

In some cases, data were missing for a country for an indicator; in these cases, missing data were replaced with a zero value. This is a conservative approach as zero values (i.e., outside the range of typical values seen in the data) inflate the total variance in the data set and thus, if anything, deflate the percent of the variance explained by PCA. Therefore, this approach avoids mistakenly attributing predictive value to principal components due to incomplete data. See **Table S3** for data sources for each variable.

### 3.2 Principal Component Analysis

The PCA was conducted on each of the three subsets described above, using the scikitlearn library ^64^. In order to avoid biasing the PCA due to large differences in magnitude and scale, each feature was centered around the mean, and scaled to unit variance prior to the analysis. Briefly, PCA applies a linear transformation to a set of *n* features to output a set of *n* orthogonal principal components which are uncorrelated and each explain a percentage of the total variance in the dataset ^65^. A link to the code for this analysis is available online at the **SSA-SARS-CoV-2-tool** (https://labmetcalf.shinyapps.io/covid19-burden-africa/).

The principal components were then analyzed for the percentage of variance explained, and compared to: (i) the number of COVID-19 tests per 100,000 population as of the end of June, 2020 (**Table S1**), (ii) the per capita GDP, and (iii) the GINI index of wealth inequality. For the GINI index, estimates from 2008-2018 were available for 45 of the 48 countries (no GINI index data were available for Eritrea, Equatorial Guinea, and Somalia) (see **Data File 1** for the year for each country for each metric).

### 3.3 PCA Results

The first two principal components from the analysis of 29 variables explain 32.6%, and 13.1% the total variance, respectively, in the dataset. Countries with higher numbers of completed SARS-CoV-2 tests reported tended to associate with an increase in principal component 1 (Pearson correlation coefficient, *r* = 0.67, *p* = 1.1e-7, **Figure S5A**). Similarly, high GDP countries seem to associate with an increase in principal component 1 (Pearson correlation coefficient, *r* = 0.80, *p* = 6.02e-12), **Figure S5B**). In contrast, countries with greater wealth inequality (as measured by the GINI index) are associated with a decrease in principal component 2 (Pearson correlation coefficient, *r* = -0.42, *p* = .0042, **Figure S5C**). Despite these correlations, a relatively low percentage of variance is explained by each principal component: for the 29 variables, 13 of the 29 principal components are required to explain 90% of the variance (**Figure S5D**). When only the access to care subset of variables is considered, the first two principal components explain 50.7% and 19.1% of the variance, respectively, and five of eight principal components are required to explain 90% of the variance. When only the comorbidities subset is considered, the first two principal components explain 27.9% and 17.8% of the variance, respectively, and nine of 14 principal components are required to explain 90% of the variance (**Figure S4D**).

### 3.4 PCA Discussion

These data suggest that inter-country variation in this dataset is not easily explained by a small number of variables. Moreover, though correlations exist between principal components and high-level explanatory variables (testing capacity, wealth), their magnitude is modest. These results highlight that dimensionality reduction is unlikely to be an effective analysis strategy for the variables considered in this study. Despite this overall finding, the PCA on the access to care subset of variables highlights that the variance in these variables is more easily explained by a small number of principal components, and hence may be more amenable to dimensionality reduction. This finding is unsurprising as, for example, the number of hospital beds per 100,000 population is likely to be directly related to the number of hospitals per 100,000 population (indeed *r* = 0.60, *p* = 5.7e-6 for SSA). In contrast, for comorbidities, the relationship between different variables is less clear. Given the low percentages of variation captured by each principal component, and the high variability between different types of variables, these results motivate a holistic approach to using these data for assessing relative SARS-CoV-2 risk across SSA.

**Figure S5.**
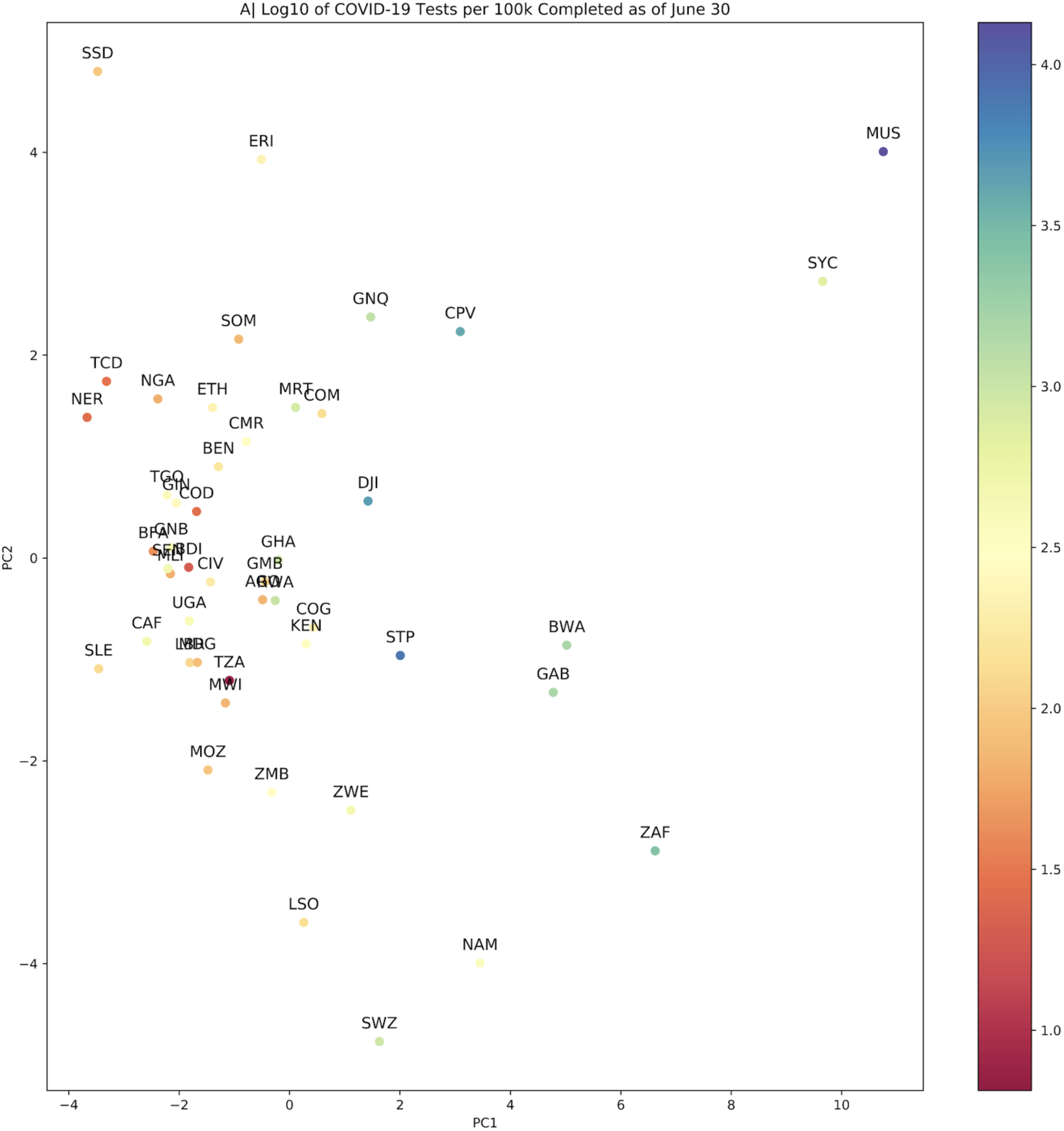
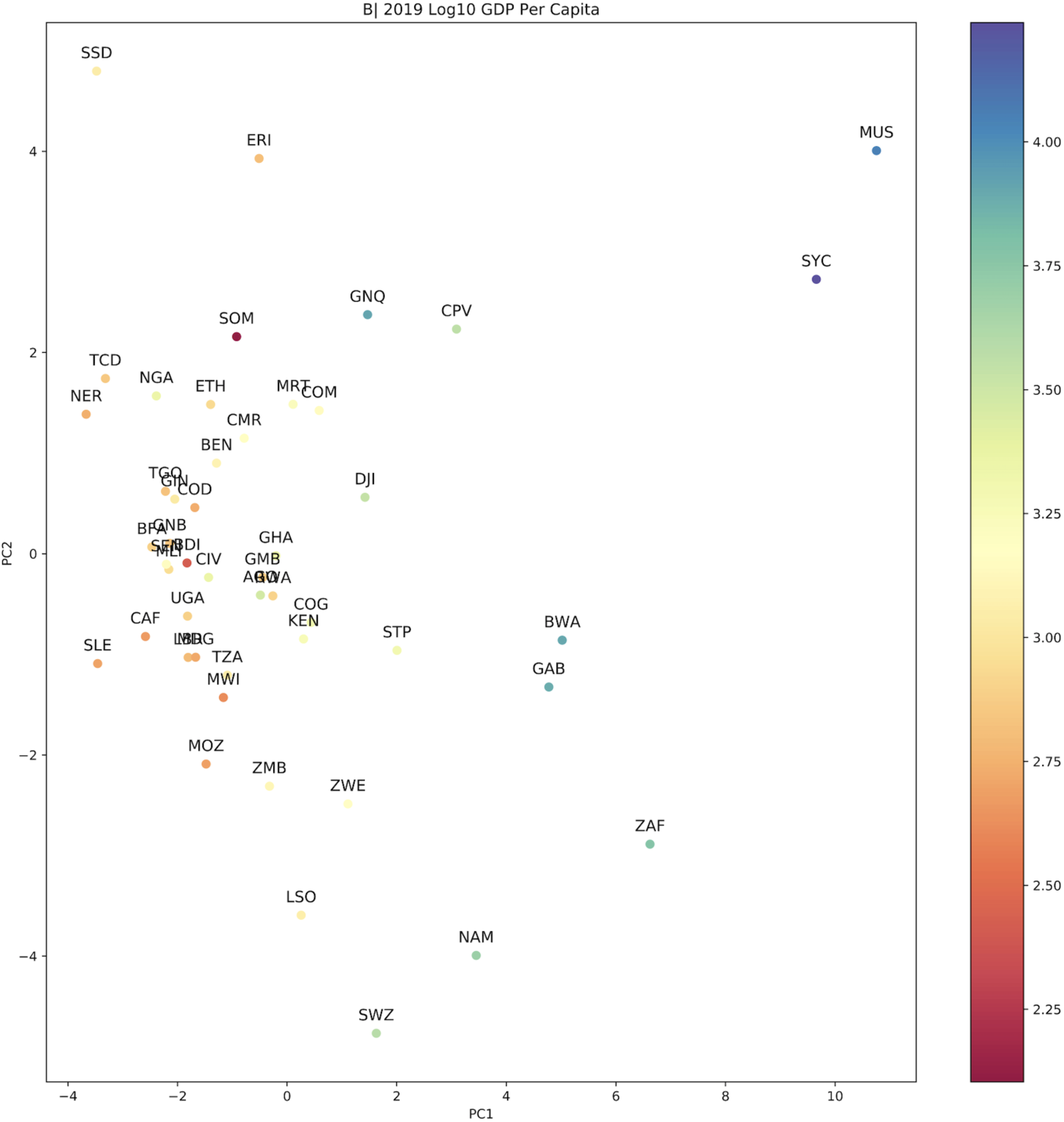
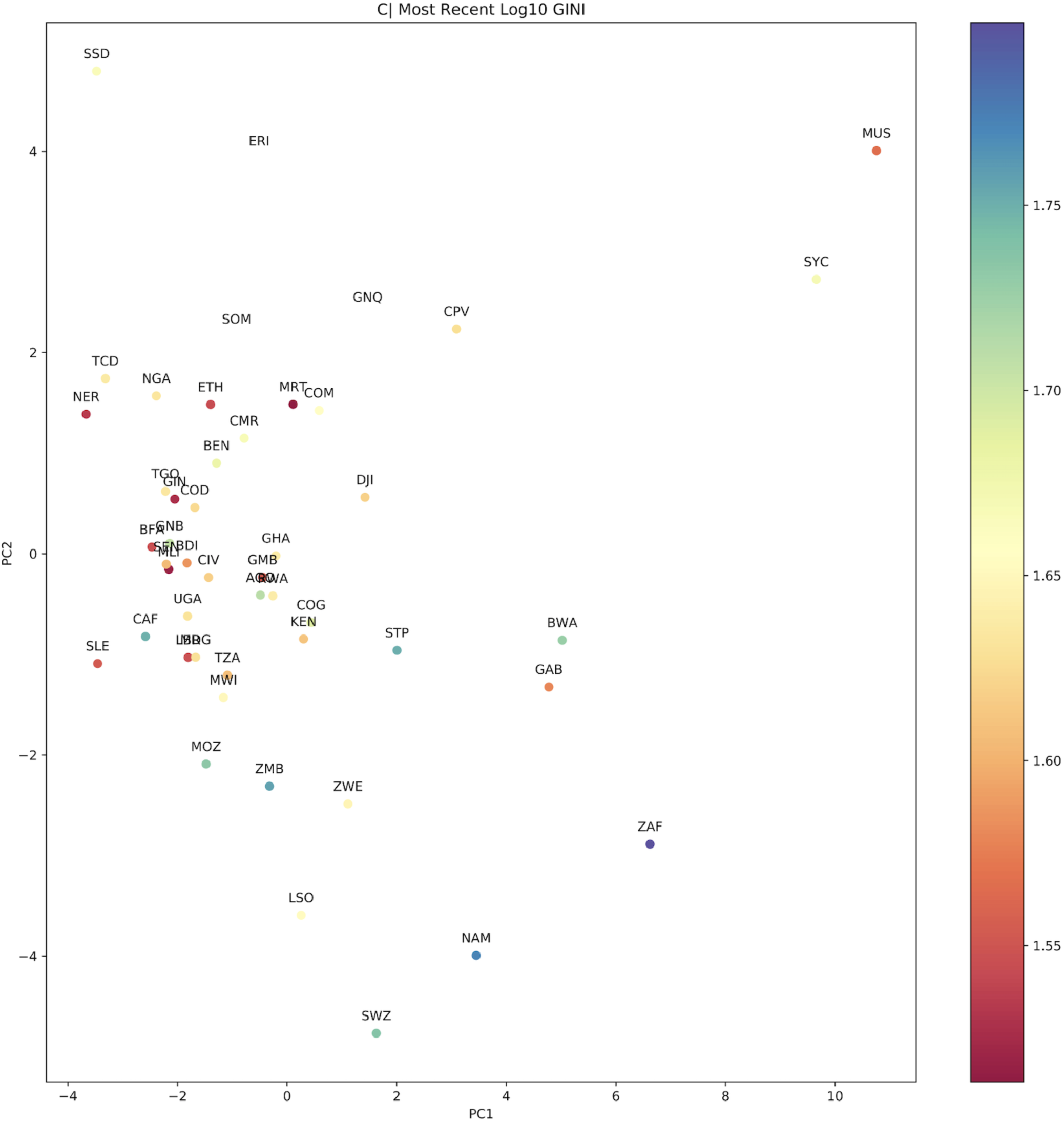
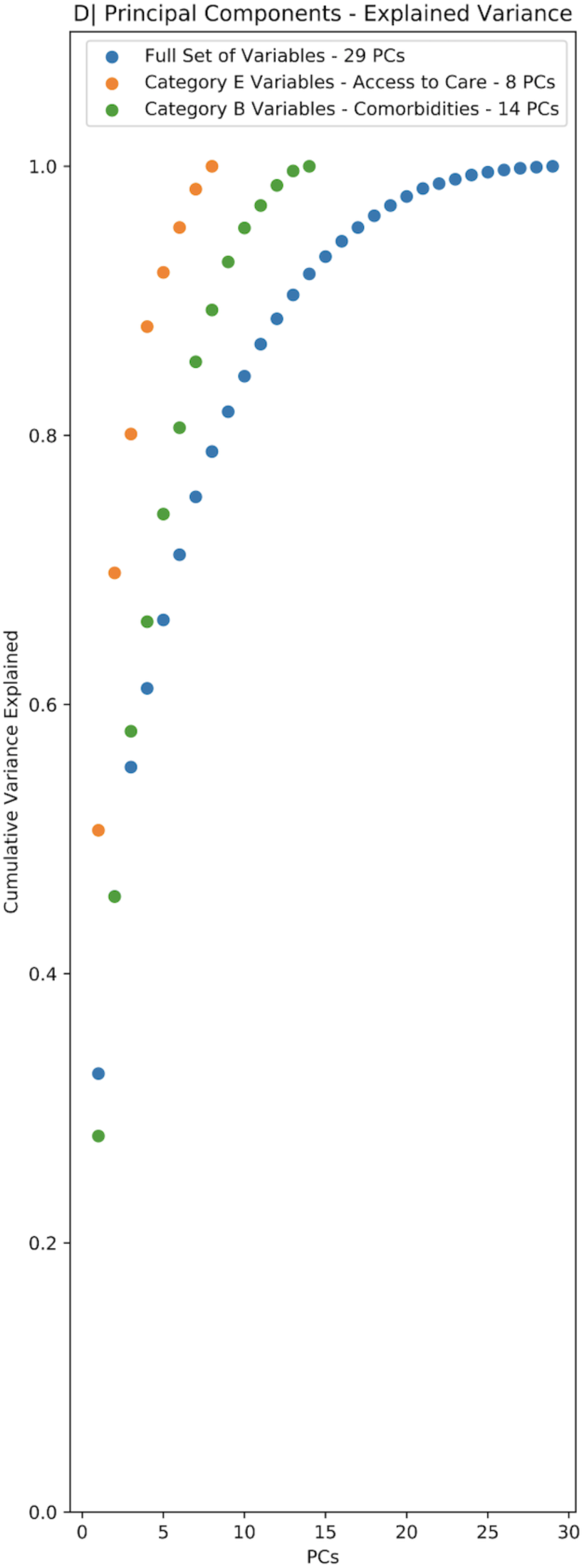
Principal Component Analysis of all variables and category specific subsets of variables. **A:** Principal Component 1 and 2, countries colored by Log10 scaled tests per 100,000 population (as of June 30, 2020) **B:** Principal Component 1 and 2, countries colored by Log10 scaled GDP per capita **C**: Principal Component 1 and 2, countries colored by the GINI index (a measure of wealth disparity) **D**: Scree plot showing the cumulative proportion of variance explained by principal component for analysis done using all variables (blue, 29 variables), comorbidity indicators (green, 14 variables, Section B in **Table S3**)), and access to care indicators (orange, 8 variables, Section E in **Table S3**)

## A4 Evaluating the burden emerging from the severity of infection outcome

### 4.1 Data sourcing: Empirical estimates of IFR

Estimates of the infection fatality ratio (*IFR*) that account for asymptomatic cases, underreporting, and delays in reporting are few, however, it is evident that *IFR* increases substantially with age ^66^. We use age-stratified estimates of *IFR* from three studies (two published ^2,4^, one preprint ^3^) that accounted for these factors in their estimation (**Table S4**).

**Table S4:**
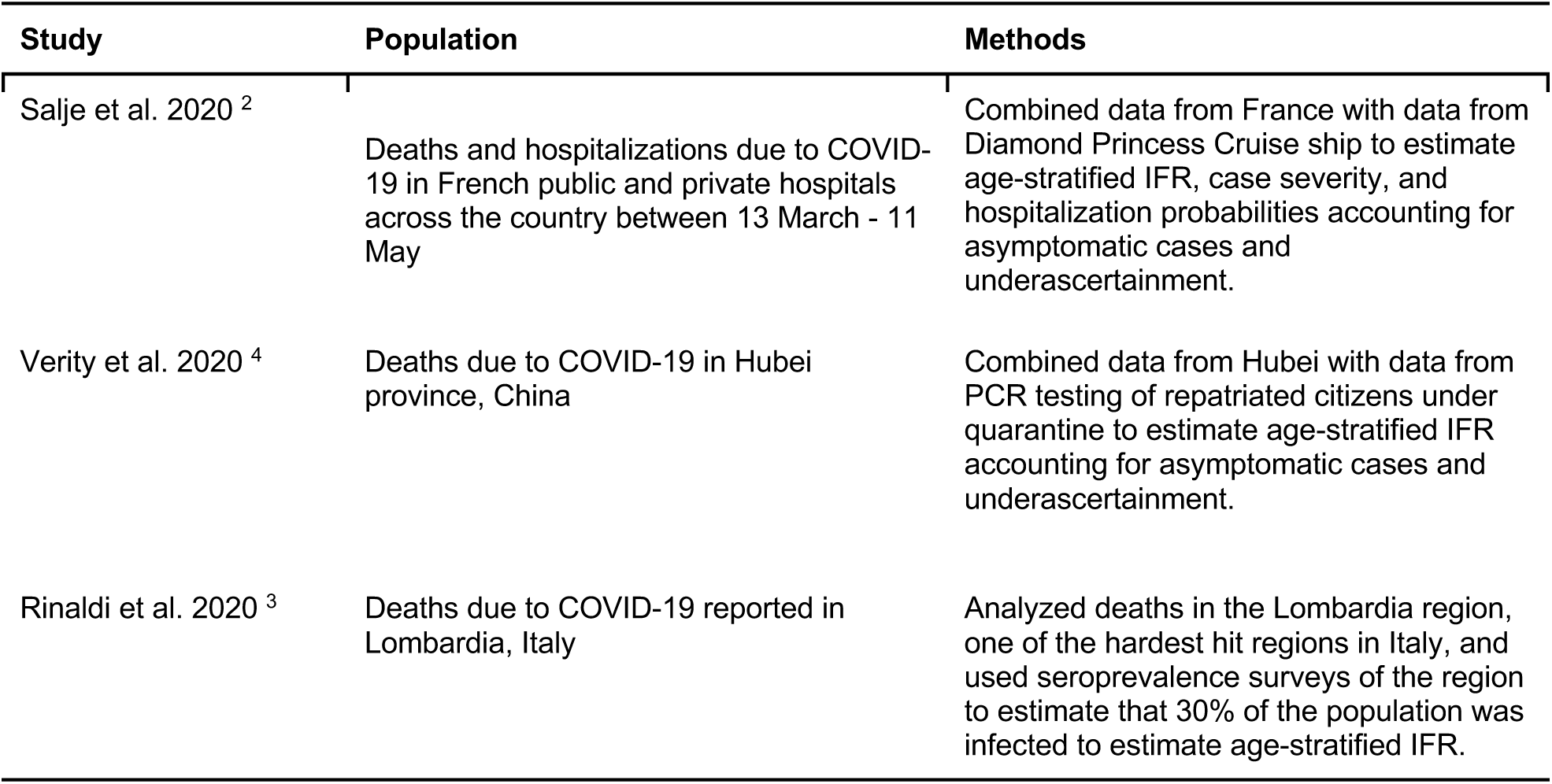
Sources of age-stratified IFR estimates

To apply these estimates to other age-stratified data with different bin ranges and generate continuous predictions of *IFR* with age, we fit the relationship between the midpoint of the age bracket and the *IFR* estimate using a generalized additive model (GAM) using the ‘mgcv’ package ^67^ in R version 4.0.2 ^68^. We use a beta distribution as the link function for IFR estimates (data distributed on [0, 1]). For the upper age bracket (80+ years), we take the upper range to be 100 years and the midpoint to be 90.

We assume a given level of cumulative infection (here 20% in each age class, i.e., a constant rate of infection among age classes) and then apply *IFRs* by age to the population structure of each country to generate estimates of burden. Age structure estimates were taken from the UNPOP (see **Table S3**) country level estimates of population in 1 year age groups (0 - 100 years of age) to generate estimates of burden.

### 4.2 Data sourcing: Comorbidities over age from IHME

Applying these *IFR* estimates to the demographic structure of SSA countries provides a baseline expectation for mortality, but depends on the assumption that mortality patterns in sub-Saharan Africa will be similar to those from where the *IFR* estimates were sourced (France, China, and Italy). Comorbidities have been shown to be an important determinant of the severity of infection outcomes (i.e., *IFR*); to assess the relative risk of comorbidities across age in SSA, estimates of comorbidity severity by age (in terms of annual deaths attributable) were obtained from the Institute for Health Metrics and Evaluation (IHME) Global Burden of Disease (GBD) study in 2017 ^69^. Data were accessed through the GBD results tool for cardiovascular disease, chronic respiratory disease (not including asthma), and diabetes, reflecting three categories of comorbidity with demonstrated associations with risk (**Table S2**). We make the assumption that higher mortality rates due to these NCDs, especially among younger age groups, is indicative of increased severity and lesser access to sufficient care for these diseases - suggesting an elevated risk for their interaction with SARS-CoV-2 as comorbidities. While there are significant uncertainties in these data, they provide the best estimates of age specific risks and have been used previously to estimate populations at risk^18^.

The comorbidity by age curves for SSA countries were compared to those for the three countries from which SARS-CoV-2 *IFR* by age estimates were sourced. Attributable mortality due to all three NCD categories is higher at age 50 in all 48 SSA countries when compared to estimates from France and Italy and for 42 of 48 SSA countries when compared to China (**Figure S5**).

Given the potential for populations in SSA to experience a differing burden of SARS-CoV-2 due to their increased severity of comorbidities in younger age groups, we explore the effects of shifting *IFRs* estimated by the GAM of *IFR* estimates from France, Italy, and China younger by 2, 5, and 10 years (**Figure 3** in main text).

**Figure S6.**
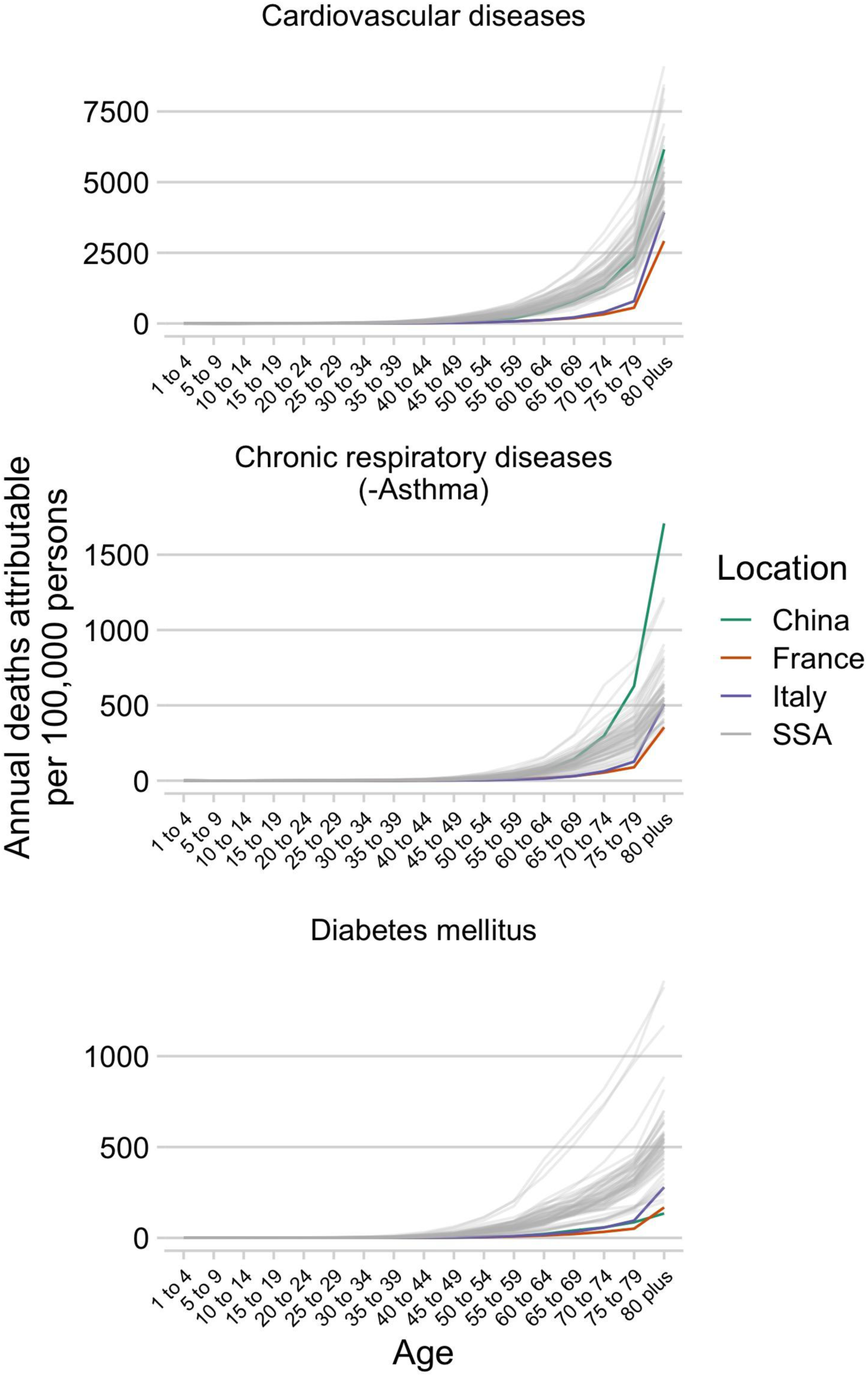
Comorbidity burden by age in sub-Saharan Africa. Estimated mortality per age group for sub-Saharan African countries (gray lines) compared to China, France, and Italy (the countries from which estimates of SARS-CoV-2 infection fatality ratios (*IFR*s) by age are available) for three NCD categories (cardiovascular diseases, chronic respiratory diseases excluding asthma, and diabetes).

## A5 International Air Travel to SSA

The number of passenger seats on flights arriving to international airports were grouped by country and month for January 2020 to April 2020 (**Table S5**) - the months when the introduction of SARS-CoV-2 to SSA countries was likely to have first occurred. The first confirmed case reported from a SSA country, per the Johns Hopkins Coronavirus Research Center was in Nigeria on February 28, 2020. By March 31, 2020, 43 of 48 SSA countries had reported SARS-CoV-2 infections and international travel was largely restricted by April. Lesotho was the last SSA country to report a confirmed SARS-CoV-2 infection (on May 13, 2020); however, given difficulties in surveillance, the first reported detections were likely delayed relative to the first importations of the virus.

The probability of importation of the virus is defined by the number of travelers from each source location each date and the probability that a traveler from that source location on that date was infectious. Due to limitations in surveillance, especially early in the SARS-CoV-2 pandemic, empirical data on infection rates among travelers is largely lacking. To account for differences in the status of the SARS-CoV-2 pandemic across source locations, and thus differences in the importation risk for travelers from those locations, we coarsely stratified travelers arriving each day into four categories based on the status of their source countries:

i. Travelers from countries with zero reported cases (i.e., although undetected transmission was possibly occurring, SARS-CoV had not yet been confirmed in the source country by that date)
ii. Those traveling from countries with more than one reported case (i.e., SARS-CoV-2 had been confirmed to be present in that source country by that date),
iii. Those traveling from countries with more than 100 reported cases (indicating community transmission was likely beginning), and
iv. Those traveling from countries with more than 1000 reported cases (indicating widespread transmission)

For determining reported case counts at source locations for travelers, no cases were reported outside of China until January 13, 2020 (the date of the first reported case in Thailand). Over January 13 to January 21, cases were then reported in Japan, South Korea, Taiwan, Hong Kong, and the United States (https://covid19.who.int/). Subsequently, counts per country were tabulated daily by the Johns Hopkins Coronavirus Resource Center ^70^ beginning January 22 (https://coronavirus.jhu.edu/map.html); we use that data from January 22 onwards and the WHO reports prior to January 22.

The number of travelers within each category arriving per month is shown in **Table S5**. This approach makes the conservative assumption that the probability a traveler is infected reflects the general countrywide infection rate of the source country at the time of travel (i.e., travelers are not more likely to be exposed than non-travelers in that source location) and does not account for complex travel itineraries (i.e., a traveler from a high risk source location transiting through a low risk source location would be grouped with other travelers from the low risk source location). Consequently, the risk for viral importation is likely systematically underestimated. However, as the relative risk for viral importation will still scale with the number of travelers, comparisons among SSA countries can be informative (e.g., SSA countries with more travelers from countries with confirmed SARS-CoV-2 transmission are at higher risk for viral importation).

**Table S5 Arrivals to SSA airports by the number of passenger seats and status of the SARS-CoV-2 pandemic at the origin at the time of travel**

(see csv file: “Table S5 International Airtravel to SSA.csv”)

Data Fields:

1. country: Name of country
2. n_airports: Number of airports with flight data
3. month: January, February, March, April 2020; or total for all 4 months
4. total_n_seats: Total number of passenger seats on arriving aircraft
5. From source with cases > 0: Number of passengers arriving from source locations with 1 or more reported SARS-CoV-2 infection by the date of travel
6. From source with cases > 100: Number of passengers arriving from source locations more than 100 reported SARS-CoV-2 infection by the date of travel
7. From source with cases > 1000: Number of passengers arriving from source locations with more than 1000 reported SARS-CoV-2 infection by the date of travel

## A6 Subnational connectivity among countries in sub-Saharan Africa

### 6.1. Indicators of subnational connectivity

To allow comparison of the relative connectivity across countries, we use the friction surface estimates provided by Weiss et al.^24^ as a relative measure of the rate of human movement between subregions of a country. For connectivity within subregions of a country (e.g., transport from a city to the rural periphery), we use as an indicator the population weighted mean travel time to the nearest urban center (i.e., population density > 1,500 per square kilometer or a density of built-up areas > 50% coincident with population > 50,000) within administrative-2 units ^63^. For some countries, estimates at administrative-2 units were unavailable (Comoros, Cape Verde, Lesotho, Mauritius, Mayotte, and Seychelles); estimates at the administrative-1 unit level were used for these cases (these were all island nations, with the exception of Lesotho).

### 6.2. Metapopulation model methods

Once SARS-CoV-2 has been introduced into a country, the degree of spread of the infection within the country will be governed by subnational mobility: the pathogen is more likely to be introduced into a location where individuals arrive more frequently than one where incoming travellers are less frequent. Large-scale consistent measures of mobility remain rare. However, recently, estimates of accessibility have been produced at a global scale ^24^. Although this is unlikely to perfectly reflect mobility within countries, especially as interventions and travel restrictions are put in place, it provides a starting point for evaluating the role of human mobility in shaping the outbreak pace across SSA. We use the inverse of a measure of the cost of travel between the centroids of administrative level 2 spatial units to describe mobility between locations (estimated by applying the costDistance function in the *gdistance* package in R to the friction surfaces supplied in ref^24^). With this, we develop a metapopulation model for each country to develop an overview of the possible range of trajectories of unchecked spread of SARS-CoV-2.

We assume that the pathogen first arrives into each country in the administrative 2 level unit with the largest population (e.g., the largest city) and the population in each administrative 2 level (of size N _*j*_) is entirely susceptible at the time of arrival. We then track spread within and between each of the administrative 2 level units of each country. Within each administrative 2 level unit, dynamics are governed by a discrete time Susceptible (S), Infected (I) and Recovered (R) model with a time-step of ∼ 1 week, which is broadly consistent with the serial interval of SARS-CoV-2. Within the spatial unit indexed *j*, with total size N _*j*_, dynamics follow:

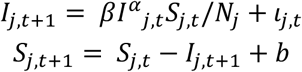

where *β*captures the magnitude of transmission over the course of one serial interval (and is set to 2.5 to approximately represent the R_0_ of SARS-CoV-2); the exponent, *α*= 0.97 is used to capture the effects of discretization^71^,l_*j,t*_ captures the introduction of new infections into site *j* at time *t*, and *b* reflects the introduction of new susceptible individuals resulting from the birth rate, set to reflect the most recent estimates for that country from the World Bank Data Bank (https://data.worldbank.org/indicator/SP.DYN.CBRT.IN).

We make the simplifying assumption that mobility linking locations *i* and *j*, denoted *c*_*i,j*_, scales with the inverse of the cost of travel between sites *i* and *j* evaluated according to the friction surface provided in ^24^. The introduction of an infected individual into location *j* is then defined by a draw from a Bernouilli distribution following:

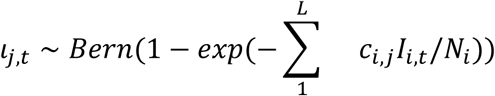

where *L* is the total number of administrative 2 units in that country, and the rate of introduction is the product of connectivity between the focal location and each other location multiplied by the proportion of population in each other location that is infected.

Some countries show rapid spread between administrative units within the country (e.g., a country with parameters that broadly reflect those available for Malawi, **Figure S7**), while in others (e.g., reflecting Madagascar), connectivity may be so low that the outbreak may be over in the administrative unit of the largest size (where it was introduced) before introductions successfully reach other poorly connected administrative units. The result is a hump shaped relationship between the fraction of the population that is infected after 5 years and the time to the first local extinction of the pathogen (**Figure S7**, right top). In countries with lower connectivity (e.g., that might resemble Madagascar), local outbreaks can go extinct rapidly before travelling very far; in other countries (e.g., that might resemble Gabon), the pathogen goes extinct rapidly because it travels rapidly and rapidly depletes susceptible individuals everywhere.

The impact of the pattern of travel between centroids is echoed by the pattern of travel within administrative districts: countries where the pathogen does not reach a large fraction of the administrative 2 units within the country in 5 years are also those where within administrative unit travel is low (**Figure S7**, right bottom).

These simulations provide a window onto qualitative patterns expected for subnational spread of the pandemic virus, but there is no clear way of calibrating the absolute rate of travel between regions of relevance for SARS-CoV-2. Thus, the time-scales of these simulations should be considered in relative, rather than absolute terms. Variation in lockdown effectiveness, or other changes in mobility for a given country may also compromise relative comparisons. Variability in case reporting complicates clarifying this (**Figure S8**).

**Figure S7.**
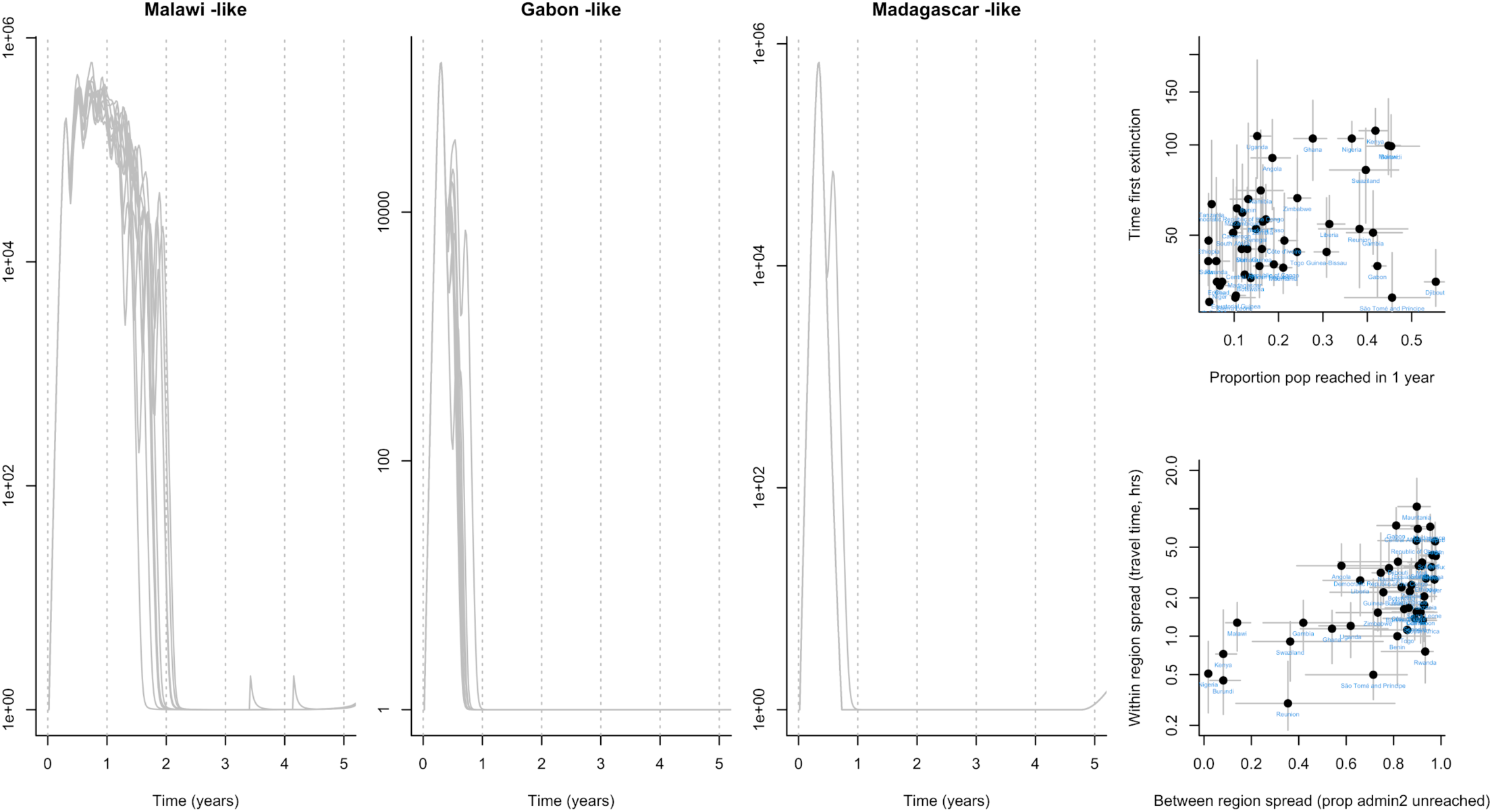
Pace of the outbreak. Each grey line on the left hand panels indicates the total infected across all administrative units in a metapopulation simulation with parameters reflecting the country indicated by the plot title, assuming interventions are constant. Increases after the first peak indicate the pathogen reaching a new administrative 2 unit. In Malawi-like settings (higher connectivity), more administrative units are reached rapidly, whereas in Madagascar-like settings (lower connectivity), a lower proportion of the administrative units are reached by a given time, as fewer introductions occur before the outbreak has burned out in the administrative 2 unit with the largest population. More generally, rapid disappearance of the outbreak (top right hand plot, y axis shows time to extinction) could either indicate rapid spread with a high proportion of the countries’ population reached (top right hand plot, x axis) or slow spread, with many administrative units unreached, and therefore remaining susceptible. The pattern of between-administrative unit travel also echoes travel time within administrative units (lower panel, right hand side, x axis indicates fraction of administrative units unreached, and upper panel indicates travel time in hours to the nearest city of 50,000 or more people).

**Figure S8.**
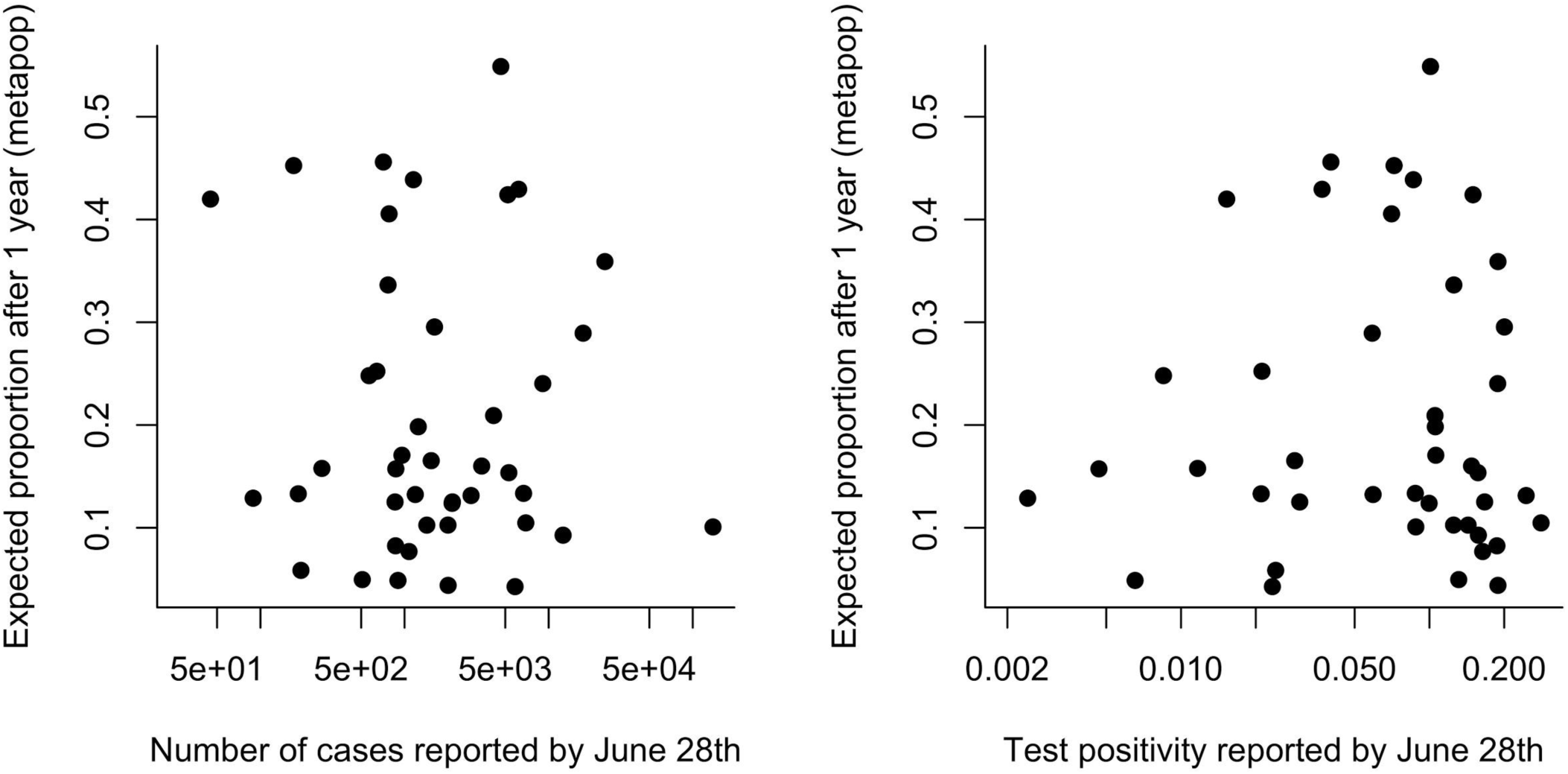
Cases and testing vs. the pace of the outbreak. The total number of confirmed cases reported by country (x axis, left, as reported for June 28th by Africa CDC) and the test positivity (x axis, right, defined as the total number of confirmed cases divided by the number of tests run, as reported by Africa CDC, likewise) show no significant relationship with the proportion of the population estimated to be infected after one year using the metapopulation simulation described in A6 (respectively, *ρ* = −0.04, *p* > 0.5, *df*, = 41 and *ρ* = 0.02, (*p*> 0.5, *df*= 41). All else equal, a positive relationship is expected; however, both uncertainty in case numbers, and uncertainty associated with the simulation might both drive the absence of a signal.

## A7 Modeling epidemic trajectories in scenarios where transmission rate depends on climate

### 7.1 Climate data sourcing: Variation in humidity in SSA

Specific humidity data for selected urban centers comes from ERA5 using an average climatology (1981-2017)^47^; we do not consider year-to-year climate variations. Selected cities (*n* = 58) were chosen to represent the major urban areas in SSA. The largest city in each SSA country was included as well as any additional cities that were among the 25 largest cities or busiest airports in SSA.

### 7.2 Methods for climate driven modelling of SARS-CoV-2

We use a climate-driven SIRS (Susceptible-Infected-Recovered-Susceptible) model to estimate epidemic trajectories (i.e., the time of peak incidence) in different cities in 2020, assuming no control measures are in place ^23,72^. The model is given by:

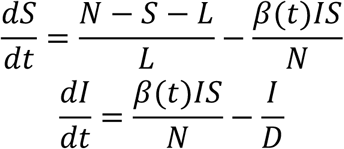

where *S* is the susceptible population, *I* is the infected population and *N* is the total population. *D* is the mean infectious period, set at 5 days following ref^23,49^. To investigate the maximum possible climate effect, we use parameters from the most climate-dependent scenario in ref^23^, based on betacoronavirus HKU1. In this scenario *L*, the duration of immunity, is found to be 66.25 weeks (i.e., greater than 1 year and such that waning immunity does not affect timing of the epidemic peak).

Transmission is governed by *β*(*t*) which is related to the basic reproduction number *R*_*0*_ by *R*_0_(*t*) = *β*(*t*)*D*. The basic reproduction number varies based on the climate and is related to specific humidity according to the equation:

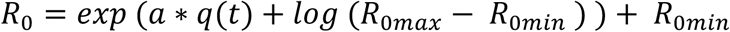

where *q*(*t*) is specific humidity ^47^ and *a* is set at -227.5 based on estimated HKU1 parameters ^23^. *R*_*0max*_ and *R*_*0min*_ are 2.5 and 1.5 respectively. We assume the same time of introduction for all cities, set at March 1^st^, 2020 (consistent with the first reported cases in SSA, **Figure S1D**)

